# Molecular Mechanism of Action of Repurposed Drugs and Traditional Chinese Medicine Used for the Treatment of Patients Infected With COVID-19: A Systematic Scoping Review

**DOI:** 10.1101/2020.04.10.20060376

**Authors:** Fui Fui Lem, Fernandes Opook, Dexter Jiunn Herng Lee, Fong Tyng Chee, Fahcina P. Lawson, Su Na Chin

## Abstract

**Background:** The emergence of COVID-19 as a pandemic has resulted in the need for urgent development of vaccines and drugs and the conduction of clinical trials to fight the outbreak. Because of the time constraints associated with the development of vaccines and effective drugs, drug repurposing and other alternative treatment methods have been used to treat patients that have been infected by the SARS-CoV-2 virus and have acquired COVID-19.

**Objective:** The objective of this systematic scoping review is to provide an overview of the molecular mechanism of action of repurposed drugs or alternative treatment medicines used to attenuate COVID-19 disease.

**Data Sources:** The research articles or grey literature, including theses, government reports, and official news online, were identified from 4 databases and 1 search engine. The full content of a total of 160 articles that fulfilled our inclusion criteria was analyzed and information about 6 drugs (ritonavir, lopinavir, oseltamivir, remdesivir, favipiravir, and chloroquine) and 4 traditional Chinese medicines (*Shuang Huang Lian Kou Fu Ye*, TCM combination of *Bu Huan Jin Zheng Qi San* and *Da Yuan Yin, Xue Bi Jing Injection* and *Qing Fei Pai Du Tang*) were extracted.

**Conclusions:** All of the repurposed drugs that have been used for the treatment of COVID-19 depend on the ability of the drug to inhibit the proliferation of the SARS-CoV-2 virus by binding to enzyme active sites, viral chain termination, or triggering of the molecular pathway, whereas traditional Chinese medicine has a pivotal role in triggering the inflammation pathway, such as the neuraminidase blocker, to fight the SARS-CoV-2 virus. This review provides an insight to experimental validation of drugs and alternative medicine used for the treatment and control of COVID-19.

## 1. Introduction

In December 2019, a novel type of viral pneumonia was discovered in Wuhan, Hubei Province, China. The International Committee of Taxonomy of Viruses has officially named the disease “COVID-19 (Corona Virus Disease 2019)” and the virus SARS-CoV-2 [1,2]. The new corona virus has rapidly spread among humans all over the world and has led to hundreds of thousands of cases within a few months. As a result, on 11^th^ March 2020 the World Health Organization (WHO) declared COVID-19 a pandemic, which is defined as “worldwide spread of a new disease”. As of 26^th^ March 2020, more than 462,684 laboratory-confirmed cases of COVID-19 have been reported globally, with the total number of deaths reaching 20,834 (4.5%) people [3]. The worldwide distribution of COVID-19 cases as of 26^th^ March 2020, 10:00 (CET) is shown in Figure 1.

**Figure 1.**
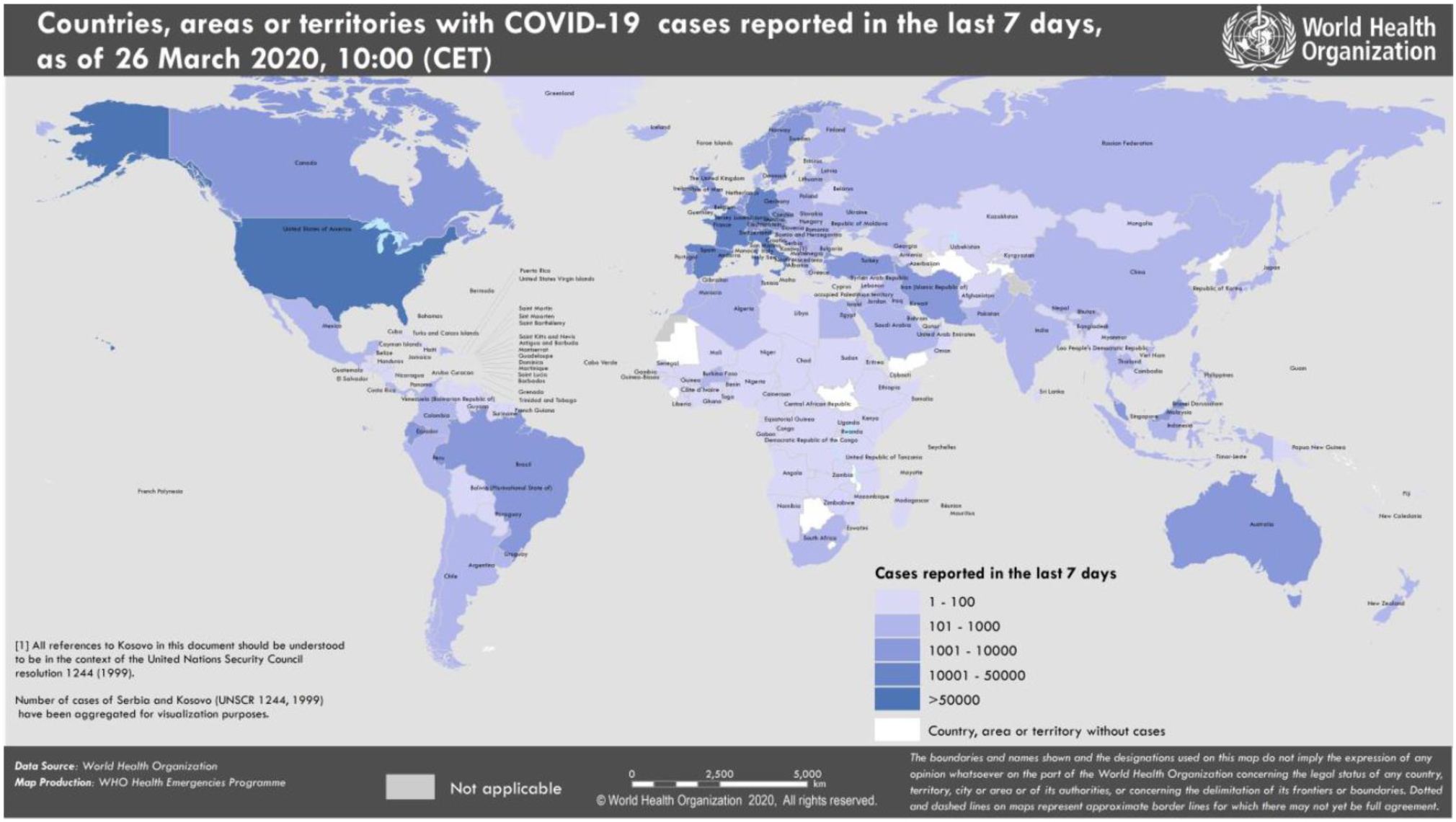
Countries, territories, or areas with reported confirmed cases of COVID-19, 26th March 2020 (Source: WHO, 2020).

CoVs are single-stranded RNA viruses that look like a crown under a microscope (coronam is the Latin word for crown) and contain spike glycoproteins on the envelope. The subfamily Orthocoronavirinae of the Coronaviridae family (order Nidovirales) is classified into four genera of CoVs: alphaCoV, betaCoV, deltaCoV, and gammaCoV. SARS-CoV-2 is categorized as a betaCoV. It has an elliptic or round and often pleomorphic form, with a diameter of 60–140 nm. Like other betaCoVs, SARS-CoV-2 is sensitive to ultraviolet rays and heat and can be effectively killed by lipid solvents. Acute respiratory illness appears to be the most common manifestation of COVID-19 infection. The spectrum of symptomatic infection ranges from mild to severe; most infections are not severe [4–6]:

- Mild clinical symptoms The initial stage occurs at the time of inoculation and early establishment of the disease. For most people, this involves an incubation period associated with mild and often non-specific symptoms such as malaise, fever, and a dry cough. During this period, SARS-CoV-2 multiplies and establishes residence in the host, primarily focusing on the respiratory system. SARS-CoV-2 binds to its target using the angiotensin-converting enzyme 2 (ACE2) receptor on human cells [7].
- Moderate clinical symptoms In the second stage of established pulmonary disease, viral multiplication and localized inflammation in the lung is the norm. During this stage, patients develop a viral pneumonia, with a cough, fever, and possibly hypoxia (defined as a PaO2/FiO2 of <300 mmHg). Imaging with a chest roentgenogram or computerized tomography reveals bilateral infiltrates or ground glass opacities. Blood tests reveal increasing lymphopenia along with transaminitis [8].
- Severe clinical symptoms Approximately 1 out of every 6 COVID-19 patients transition into the third and most severe stage of the illness, which manifests as an extrapulmonary systemic hyperinflammation syndrome. In this stage, markers of systemic inflammation appear to be elevated. COVID-19 infection results in a decrease in helper, suppressor, and regulatory T cell counts [9]. In this stage, shock, vasoplegia, respiratory failure, and even cardiopulmonary collapse are discernible. Systemic organ involvement, even myocarditis, can manifest during this stage.

The latest trend shows that human-to-human spread is the main mode of transmission, which is believed to happen through respiratory droplets from sneezing and coughing. Aerosol transmission is also possible in closed areas. Infection might also happen if someone touches a contaminated surface and then touches their own eyes, nose, or mouth [10–12]. Although symptomatic people are the most frequent source of COVID-19 spread, the possibility of transmission before symptoms develop, or even from individuals who remain asymptomatic, cannot be excluded. Moreover, the period during which an individual with COVID-19 is infectious is uncertain. The duration of viral shedding is also variable [12–15]. Data suggests that the use of social distancing is the best way to control this pandemic. Several countries have taken measures such as mobility restrictions, drastic social distancing, school closures, and travel bans, which could significantly disrupt economic and social stability.

At this moment, the therapeutic approaches to handle COVID-19 are only supportive. There is neither a vaccine to prevent infections nor clinically approved antiviral drugs to treat COVID-19; therefore, identifying drug treatment options as soon as possible is critical for responding to the pandemic. Clinical trials for vaccines are currently underway. Potential vaccines have been administered to volunteers in a phase 1 safety trial by the USA government; however, the efficacy, such as how long immunity will last or if people might become infected even if they possess a high level of antibodies against SARS-CoV-2, will not become clear for at least one year after injection [16]. Furthermore, the safety of the developed vaccines is unknown because laboratory tests are being conducted in parallel to clinical trial phase 1 owing to the emergence of COVID-19 as a pandemic. The unknown efficacy and safety of the vaccines used might cause disease enhancement, by which vaccinated subjects might develop an even more severe form of disease than the subjects that have not been vaccinated, which has been shown in studies of SARS vaccines, in which vaccinated ferrets developed damaging inflammation in their livers after they were infected with the virus [17]. Until now, no drugs have been successfully developed for the control of COVID-19 [18]; however, numerous effort are underway worldwide, and particularly in China [19]. Therefore, drug repurposing and the use of existing alternative medicine have been used as effective methods for the treatment of patients with COVID-19; however previous review articles have not focused on the molecular mechanism of action of these drugs. In this comprehensive review, we reviewed the existing drugs or alternative treatment methods that have been recently used and discuss the mode of action from a molecular mechanism perspective to attenuate COVID-19 in the human system.

## 2. Results

This section may be divided by subheadings. It should provide a concise and precise description of the experimental results, their interpretation as well as the experimental conclusions that can be drawn. The primary search identified 8074 published and unpublished papers, of which 2132 were from PubMed, 2775 from ScienceDirect, 720 from google scholar, 1190 from Semantic Scholar, and 1257 from Google search engine. 3798 duplicates were excluded, leaving 4276 articles for screening by title analysis. 841 eligible published and unpublished papers were identified after excluding 3435 papers that had a different theme from this systematic scoping review. In conclusion, the content of 841 papers was fully analyzed, of which 681 were excluded according to our exclusion criteria, which was medication that was not administrated as treatment for COVID-19 before 18 March 2020. We divided the drug classes into different subclasses and individual drugs to identify differences within a drug class. Repurposed drugs used for treatment of COVID-19 consisted of 6 drugs (ritonavir, lopinavir, oseltamivir, remdesivir, favipiravir and chloroquine) and 4 TCMs (*Shuang Huang Lian Kou Fu Ye*, TCM combination of *Bu Huan Jin Zheng Qi San* and *Da Yuan Yin*, *Xue Bi Jing Injection* and *Qing Fei Pai Du Tang*). The 160 articles in this review consists of 6 guidelines, 11 clinical trial registries, 11 from official news, 42 *in vitro* and *in vivo* studies, 51 reports presenting *in vitro* and *in vivo* outcomes, and 39 clinical findings and treatment efficacy. A flowchart illustrating the progressive study selection and numbers at each stage is shown in Figure 2.

**Figure 2.**
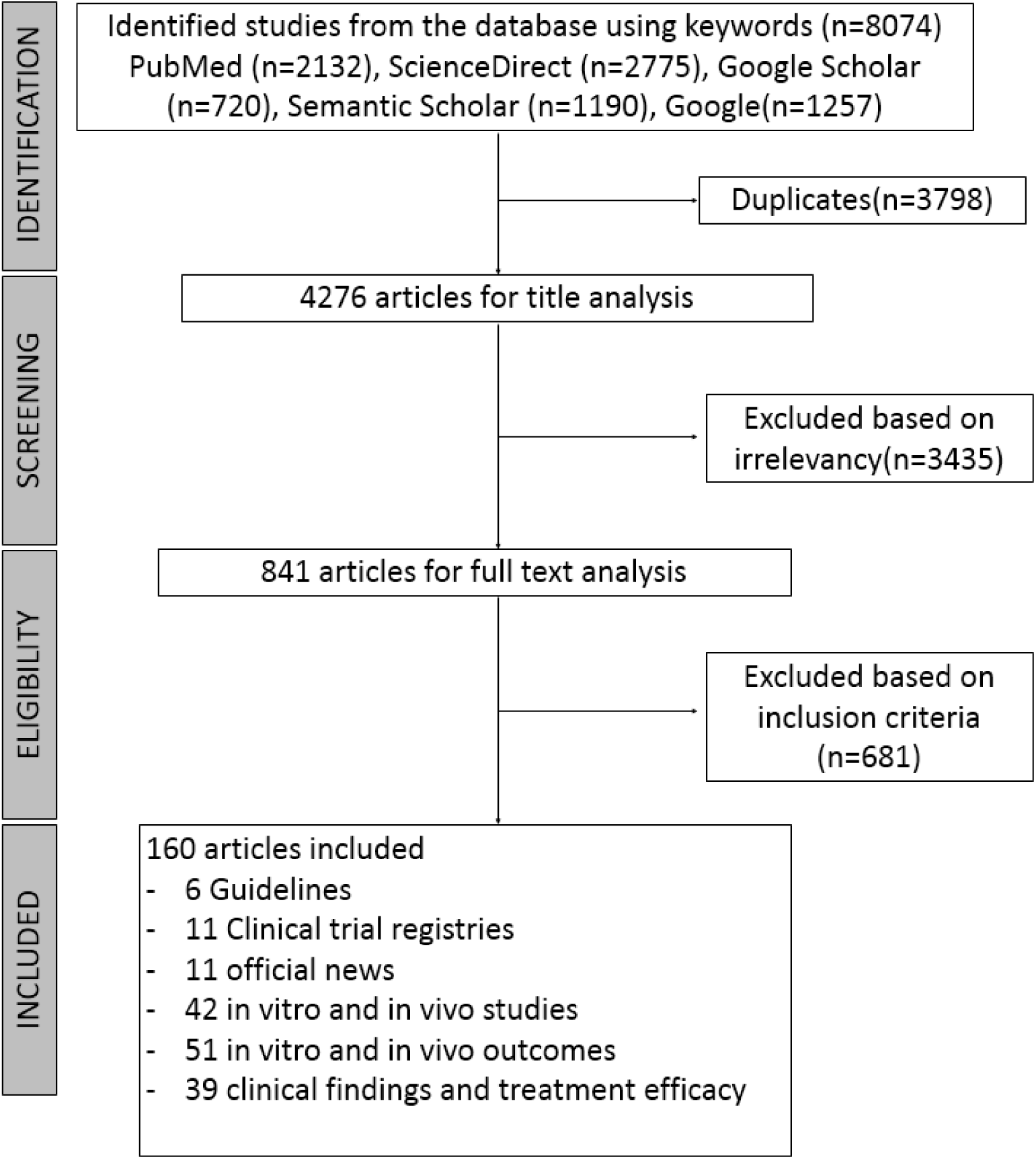
Flow diagram of the systematic search according to the guidelines for Preferred Reporting Items for Systematic Reviews (PRISMA).

## 3. Discussion

In this systematic scoping review, we provide an overview of previous studies in which existing drugs or TCMs have been repurposed for the treatment of COVID-19 and discuss the initial mechanism of action of these drugs from a molecular perspective for the treatment of coronaviruses or other viruses. The mechanism of action of these drugs give risen for its as medication used to treat COVID-19 since the outbreak of the disease in December 2019 because the development of vaccines and efficacy of clinical trial is time consuming. Recently, WHO reported the launch of a multiarm and multicountry clinical trial of 4 drugs, remdesivir, lopinavir, and ritonavir (kaletra), kaletra and interferon beta and chloroquine on 18 March 2020 [20]. Even though our systematic scoping review method of searching the literature covered medication of repurposing drugs used until 18 March 2020, most of our findings are relevant and still up to date as 4 of the drugs that have been tested in a clinical trial launched by WHO known as SOLIDARITY are covered in this review.

### 3.1. Repurposed drugs as drug candidates in the SOLIDARITY trial

#### 3.1.3. HIV protease inhibitors and anti-influenza drug as a starting point for COVID-19 treatment

During the 1990s, AbbVie Inc patented two antiretroviral protease inhibitors, ritonavir and lopinavir [21]. Ritonavir, also known as norvir, is derived from L-valine, whereas lopinavir is structurally related to ritonavir, as illustrated in Figure 3 [22]. Initially, they were developed as a standalone antiviral agent for the treatment of HIV infections, however they are combined to obtain a more efficient drug response and sold under the brand name Kaletra [23,24].

**Figure 3.**
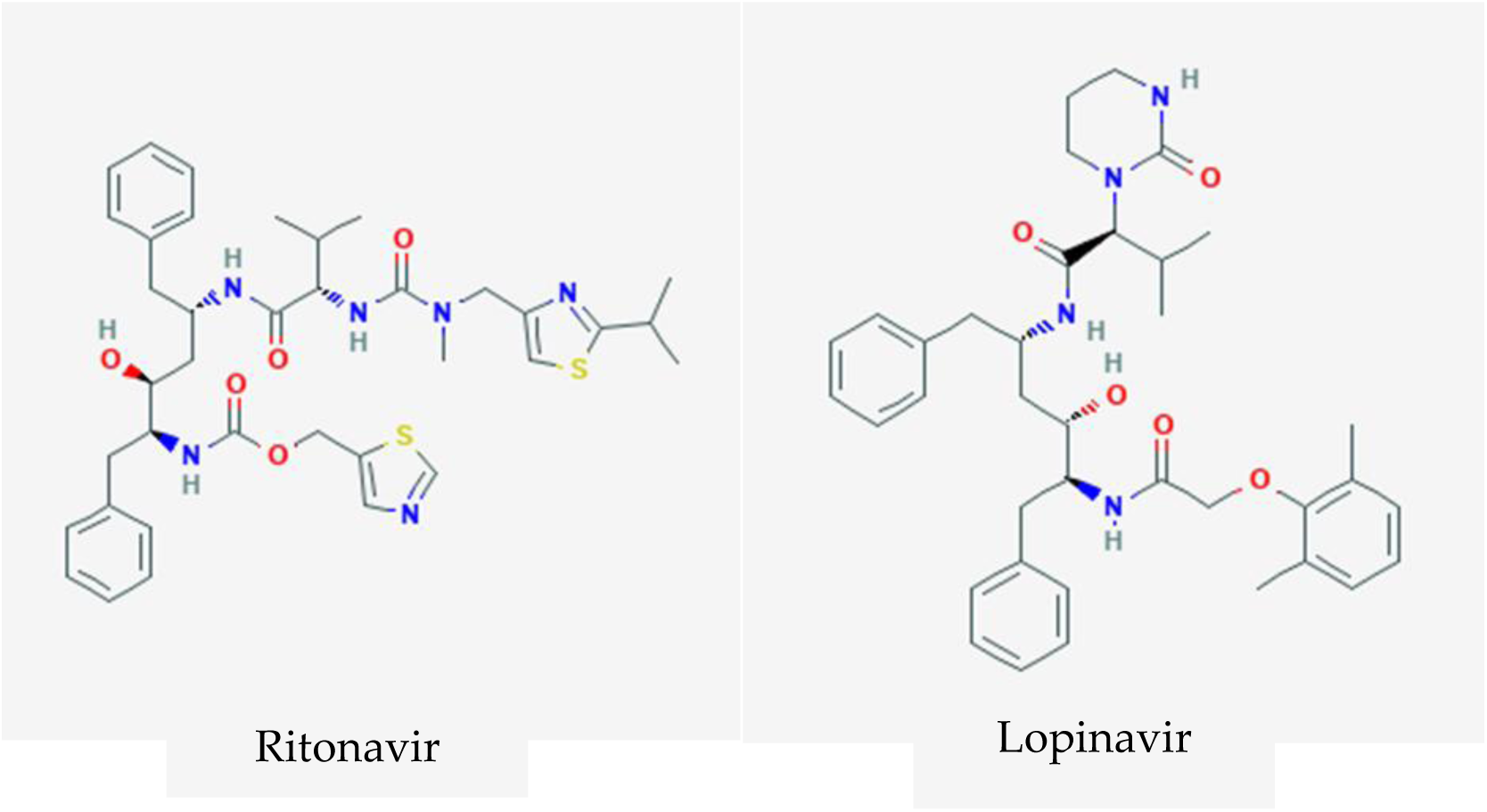
Chemical structure of HIV protease inhibitors ritonavir [25] and lopinavir [26]. The combination of ritonavir and lopinavir, i.e., kaletra, was hypothesized to function through binding on the active site of the SARS-CoV 3CL^pro^ enzyme.

Prior to the discovery and synthesis of lopinavir, ritonavir was used for the treatment of HIV-1 and HIV-2 infections through reversible inhibition of the HIV proteases by blocking access to the proteases’ active site, thus preventing the processing of the HIV Gag and Gag-Pol polyproteins [27]. This results in the formation of immature HIV particles that are not infectious. However, the extremely high mutation rate of HIV-1 *in vivo* [28] has given rise to a strain of HIV-1 that is resistant to ritonavir. The rise of ritonavir-resistant HIV strains has led the development of more effective drugs for combating HIV infections, one of which is lopinavir. Lopinavir, being a protease inhibitor, functions similarly to ritonavir in that it reversibly binds to the HIV protease’s active site and prevents the Gag and Gag-Pol polyproteins from being processed [29]. Compared with ritonavir, a lower amount of lopinavir is required to achieve EC_50_ (17 nM versus 25 nM) [30]. Although it is more effective (in terms of concentration required to achieve EC_50_), *in vitro* experiments with human liver microsomes have revealed that lopinavir undergoes oxidative metabolism by the cytochrome P450 3A4 enzyme [31], thus reducing the bioavailability of lopinavir. To combat this, lopinavir is administered together with ritonavir, which can inhibit P450 3A4 enzyme activity. The high rate of mutation of HIV has given rise to drug-resistant (against lopinavir/ritonavir) HIV strains, with lopinavir/ritonavir acting as the selective pressure. By combining these drugs together, it provide better efficacy as HIV protease inhibitors.

These drugs were the first repurposed drug that were used to treat patients diagnosed with COVID-19 [32]. The administration of ritonavir and lopinavir for the treatment of COVID-19 might be owing to the similarity of the genome sequence to SARS-CoV approximately 79.6% and originated from the same genus as SARS-CoV-2, SARS-CoV, and MERS-CoV (*Betacoronavirus*) [4]. Therefore, therapies and drugs developed for the treatment of SARS-CoV and MERS-CoV could be used for the development of COVID-19 therapeutics and assumed that the mechanism of SARS-CoV-2 is similar to its family members [22]. Lopinavir/ritonavir have been administered to patients diagnosed with moderate stage COVID-19, which is the second stage of established pulmonary disease and viral multiplication [8]. The successful treatment of COVID-19 patients with lopinavir and ritonavir have also been reported in India (an Italian couple consisting of a 69 year old male and a 70 year old female) [33] and Spain (62 year old male) [34].

A molecular dynamics simulation suggested that lopinavir and ritonavir are able to inhibit the SARS-CoV 3CL^pro^ enzyme (required for the processing of the SARS-CoV polyprotein containing the replicase enzyme) by binding to the enzyme’s active site, with neither of them having a higher binding affinity than the other [35]. However, a binding analysis showed that half of the lopinavir was left outside of the catalytic site and one of ritonavir’s benzene side chains might be too long to perfectly fit the substrate binding pocket. This would lead to ritonavir and lopinavir having poor efficacy, and this is reflected in weak *in vitro* activity against SARS-CoV [36]. Another study suggests that both lopinavir and ritonavir have no effect on SARS-CoV replication, with nelfinavir being the only inhibitor that has an effect on SARS-CoV replication [37]. Despite not having any effect on SARS-CoV replication, lopinavir does have antiviral activity against SARS-CoV, with one study suggesting lopinavir has a synergistic effect when used with ribavirin [38]. A study conducted on lopinavir’s and ritonavir’s effectiveness against MERS-CoV showed that although lopinavir and ritonavir have antiviral activity against MERS-CoV, their effectiveness is lower than interferon beta and remdesivir [39].

A recent COVID-19 study revealed that SARS-CoV-2 utilizes the same cell entry method used by SAR-CoV, namely, relying on the ACE2 receptor and priming of the spike protein by TMPRSS2 [40]. Clinically proven protease inhibitors for the ACE2 receptor and TMPRSS2 are already available in the form of camostat mesylate and E-64d. The same study also mentioned that antibodies produced against SARS-CoV can be used to combat SARS-CoV-2, albeit at a lower efficiency. Antibodies recovered from recovered COVID-19 patients could be used to combat COVID-19, although only as a preventive measure or in the early stages of the infection [41]. However, this method was successfully used to treat seriously ill patients in China, with patients showing improvement within 24 hours [42].

#### 3.1.2. Influenza drug administered in combination with a HIV protease inhibitor provide better outcome

Oseltamivir is another synthetic prodrug (Figure 4) used for the treatment for COVID-19. It is capable of inhibiting the neuraminidase enzymes on the surface of influenza virus particles. This drug is manufactured by Hoffmann-La Roche Inc, with the patent held by Gilead Sciences Inc and sold under the brand name of Tamiflu [24]. This drug is used to treat influenza A and influenza B infections, as well as for the prevention of influenza infections [43]. Oseltamivir is administered orally in its prodrug form, oseltamivir phosphate, for the treatment and prophylaxis of influenza A and influenza B infections. Oseltamivir phosphate is readily absorbed in the gastrointestinal tract, after which it is converted by hepatic esterases into its active form, oseltamivir carboxylase, a competitive inhibitor to neuraminidase found in influenza A and influenza B [24]. It reversibly binds to the active site of the neuraminidase, preventing the neuraminidase from cleaving the sialic acid residues [44] found on the surface of the host cell. This prevents the entry of the virus into uninfected cells, the release of newly formed virions from the infected cells, and reduces both viral shedding and infectivity of the virus [24]. Oseltamivir is eliminated from the body in the urine, and *in vitro* studies have shown that both oseltamivir phosphate and oseltamivir carboxylate is not susceptible to metabolism by the cytochrome P450 enzymes [45]. Although it can be used to prevent influenza infections, consumption of oseltamivir is not a substitute for influenza vaccination [45]. The common side effects of oseltamivir are nausea and vomiting, ranging from mild to moderate severity. These side effects can be mitigated by consuming oseltamivir with food [46].

**Figure 4.**
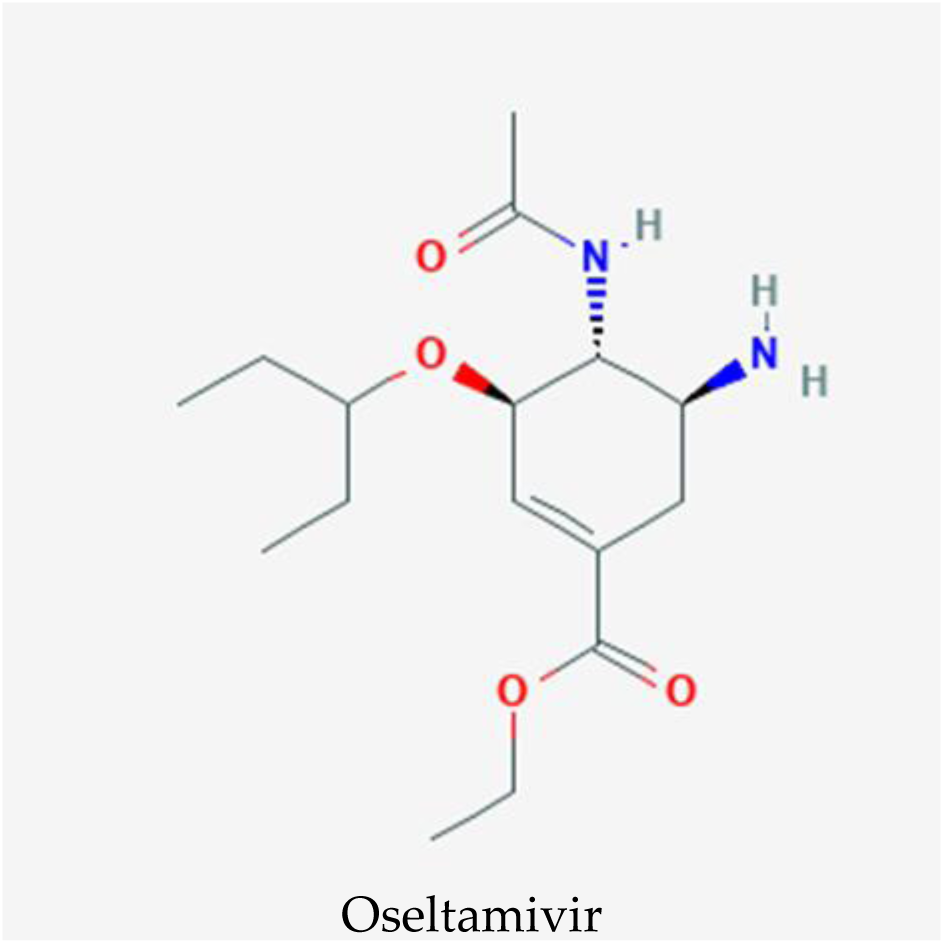
Oseltamivir, a synthetic prodrug for treatment of the Influenza virus. Oseltamivir was designed to have a similar structure to that of sialic acid, intended as a substrate in host cell membranes for neuraminidase, an enzyme on the influenza virus protein coating to hydrolyzes on to let new viral particles to infect other cells [47].

On 18 February 2020, a 74-year-old Chinese woman with COVID-19 in Thailand was treated at Rajvitjhi Hospital for COVID-19-related pneumonia with a cocktail of HIV and flu drugs [32]. The patient was first administered ritonavir and lopinavir for 5 days. After failing to show signs of recovery, oseltamivir was administered to relieve the cough and fever symptoms and reduce the severity of these symptoms in the second stage of COVID-19 infection [8]. This led to a marked improvement in her pneumonia condition in 8 – 12 hours. She tested negative for the COVID-19 after 48 hours. The drug cocktail was administered for the next 10 days, and subsequent tests for COVID-19 over the next 20 days gave negative results. However, the synergistic effect of the combination of these drugs is unclear because oseltamivir does not inhibit SARS-CoV [48] and MERS-CoV like lopinavir and ritonavir [49]. The repurposing of ritonavir, lopinavir, and oseltamivir for the treatment of COVID-19 is currently being studied by companies such as AbbVie Inc [33].

#### 3.1.3. Chain termination of viral RNA synthesis by nucleotide analogues to combat COVID-19

Remdesivir (RDV; development code GS-5734) is a 1’-cyano-substituted adenosine nucleotide analogue prodrug that was developed by Gilead Sciences in 2017 as a treatment for Ebola virus infection (Figure 5) [50]. Several studies have revealed that it has broad-spectrum antiviral activity against RNA viruses such Ebola virus (EBOV), MERS-CoV, SARS-CoV, Marburg, respiratory syncytial virus (RSV), Nipah virus (NiV), and Hendra virus [51–53]. Remdesivir has been used on a compassionate basis in several unique cases of Ebola virus disease such as to treat a newborn baby with congenital EBOV infection in combination with ZMapp and a buffy coat transfusion from an Ebola survivor [54]. Remdesivir has also been included in a larger scale therapeutic study for Ebola virus disease in the Democratic Republic of Congo. The study enrolled 681 patients from November 20, 2018, to August 9, 2019. However, the study showed a greater survival rate with monoclonal antibodies (REGN-EB3 or mAb114) treatment compared with remdesivir and ZMapp [55].

**Figure 5.**
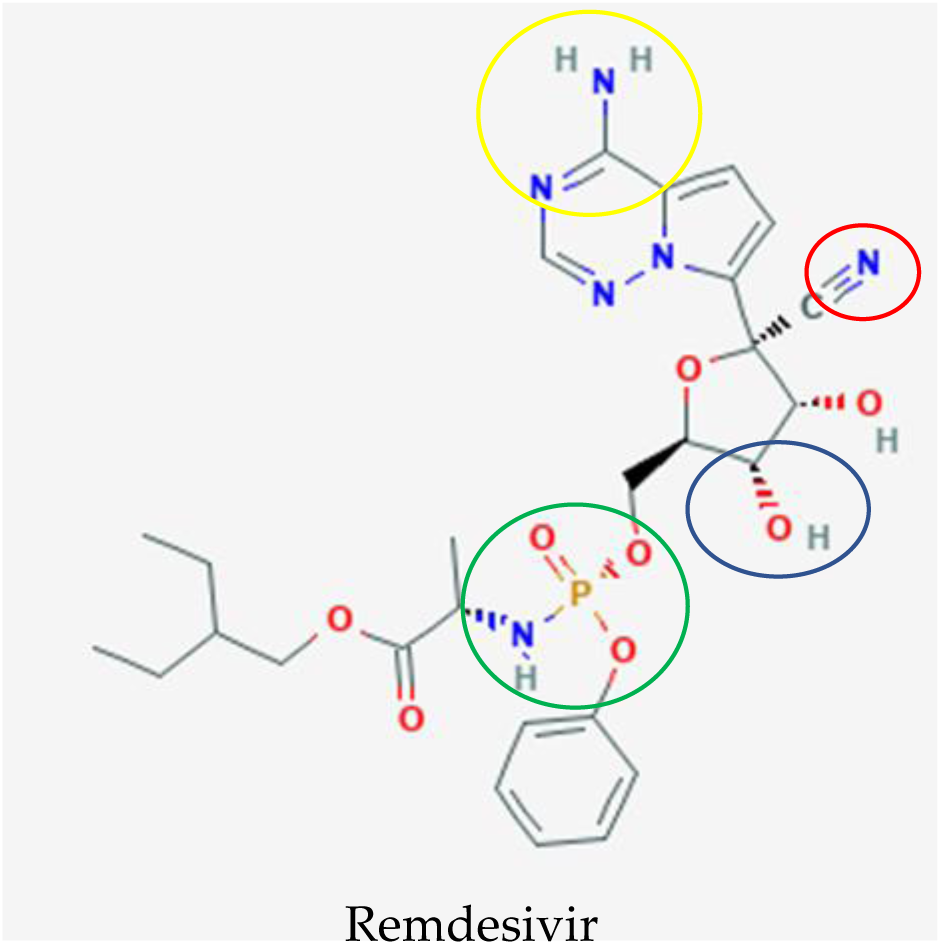
Potential drug candidate to treat COVID-19 manufactured by Gilead Sciences to treat COVID-19, remdesivir [56]. The additional of 3ʺ hydroxyl group (blue circle) in remdesivir inhibits the replication of the MERS RNA strand after a few cycles of nucleotide addition, which shields this antiviral drug from removal by coronavirus proofreading enzymes that abolish nucleotide analogs whereas the nitrogens (yellow circle) function as a proton donor and acceptor for a hydrogen bond to a uracil base as an identical binding site of double stranded RNA in adenosine. The phosphate group (green circle) is created as a protecting group or McGuigan ProTide to transport this antiviral compound into cells through phosphorylation, which is identical to a normal nucleotide triphosphate and recognized by polymerases in the cell. The important and unique part for this nucleotide analog are the addition of the 1ʹ cyano group (red circle) to eliminate the side effect of blockage of the mitochondrial RNA polymerase exhibited in mice.

Remdesivir blocks the proliferation of the RNA viruses in the host body through pre-mature termination of viral RNA transcription [50,53,57]. Remdesivir is metabolized to an active nucleoside triphosphate and works as an incorporation competitor with adenosine triphosphate. The mechanism interferes with the action of viral RNA polymerase, causing delayed chain termination and leading to decreased viral RNA production [50,53,57]. Based on an *in vitro* test utilizing primary human lung epithelial cell cultures, remdesivir was potently antiviral against Bat-CoVs, zoonotic Bat-CoVs, SARS-CoV, MERS-CoV and circulating contemporary human-CoVs [52,58,59]. In comparison with other antiviral drugs such as lopinavir, ritonavir, and interferon beta, remdesivir displayed superior antiviral activity *in vitro* using Calu-3 cells with MERS-nanoluciferase and *in vivo* in a transgenic mouse with a humanized MERS-CoV receptor (dipeptidyl peptidase 4, hDPP4) (Sheahan, 2020). The study stated that both prophylactic and therapeutic remdesivir improve pulmonary function and reduce lung viral loads and severe lung pathology. The data prompted an efficacy test of prophylactic and therapeutic remdesivir treatment in a nonhuman primate model of MERS-CoV infection, the rhesus macaque [60]. The induction of clinical disease by MERS-CoV was completely prevented with inhibition of MERS-CoV formation and lung lesion formation after prophylactic treatment initiated 24 h prior to inoculation. The data also strongly suggested a clinical benefit for therapeutic treatment with reduced virus replication and severity of lung lesions.

The improvement of severe lung pathology from *in vitro* and *in vivo* study by Sheahan [61] might help to improve the illness of patients in severe category of COVID-19, which manifests as an extra-pulmonary systemic hyperinflammation syndrome [9] and make remdesivir high potential as drug candidates that would be effective for combating COVID-19. A recent study by Wang *et al*. [62] reported chloroquine and remdesivir effectively inhibit SARS-CoV-2 infection in Vero E6 cells. This drug was even used to treat the first US patient from Washington, who was diagnosed with COVID-19 owing to a pneumonia condition [63]. Because the use of remdesivir dramatically improved the patient’s condition, a phase III clinical trial in China, United States, Hong Kong, Republic of Korea, Singapore, and France has been approved to evaluate the efficacy and safety of the drug in patients with COVID-19 (U.S National Library of Medicine, 2020) [64]. Patients in the experimental group received an initial dose of 200 mg of remdesivir followed by a daily dose of 100 mg of remdesivir through intravenous infusion in addition to standard of care therapy, whereas patients in the control group received standard of care therapy and the same dose of a RDV placebo according to the clinical trial data from U.S National Library of Medicine (2020)[64]. The clinical trial is still underway and no broad conclusions have been made about the efficacy and safety of the drug.

Favipiravir is another nucleotide analogue that has been used to treat COVID-19. Favipiravir (FPV; T-705) was developed by Toyama Chemicals in 2002 and has been approved in Japan for the treatment of influenza virus infections (Figure 6) [65,66]. Favipiravir has demonstrated antiviral activity against a wide range of RNA viruses including norovirus [67], Zika virus [68], foot-and-mouth disease virus (FMDV) [69], Rabies [70] and Ebola virus [71,72].

**Figure 6.**
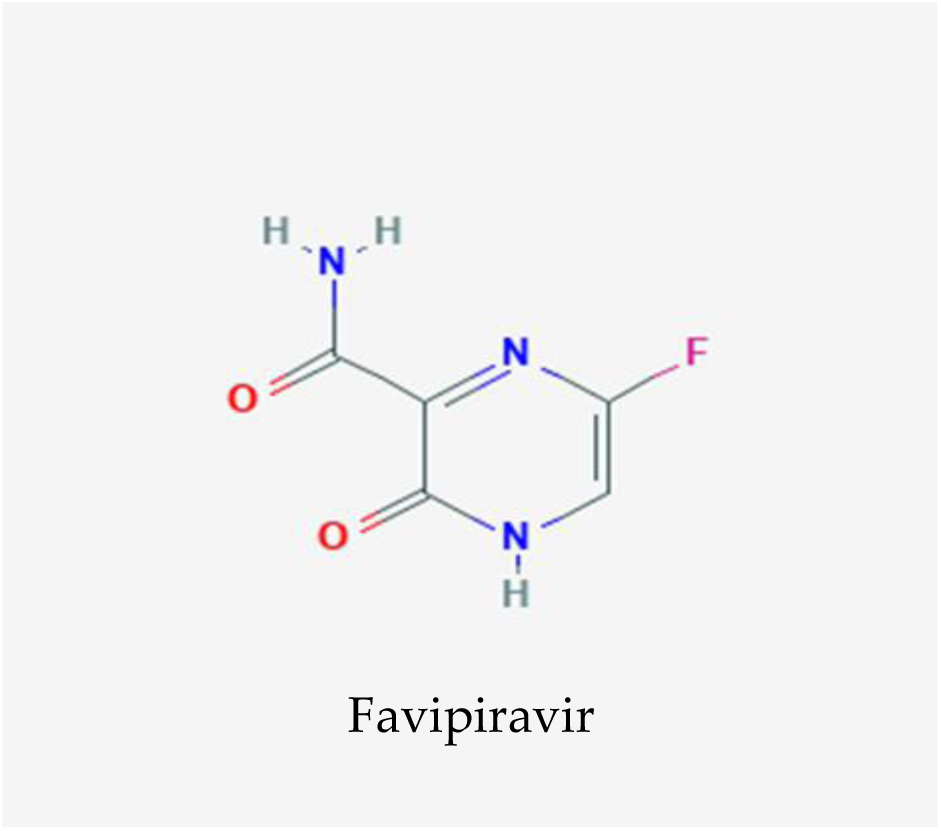
Favipiravir, nucleotide analogue manufactured by Toyama Chemicals for inhibiting a wide range of RNA viruses proliferation [73]. Favipiravir acted as an inhibitor to alter the drug binding pocket in RNA-dependent RNA polymerase of SARS-CoV-2

Like remdesivir, favipiravir inhibits viral RNA synthesis through chain termination [74,75]. Favipiravir is metabolized into ribofuranosyl 5’-triphosphate (RTP) and incorporated in the growing RNA strand. The incorporation of a single molecule of favipiravir-RTP partially prevented the extension of an RNA strand and double incorporation of favipiravir-RTP molecules completely blocked the further extension. This mechanism efficiently inhibits the viral RNA-dependent RNA polymerase function [66,76–80]. Other studies have also reported induced lethal mutagenesis during influenza A H1N1 virus and Hepatitis C virus infections and reduced viral titre *in vitro* [81,82] via favipiravir. An *in vitro* test of EBOV-infected Vero E6 cells concluded the ability of favipiravir to suppress EBOV replication [71]. Some studies have also reported the therapeutic efficacy of favipiravir to clear the virus in small animal models (transgenic mice), in which favipiravir reduces viremia and prevents the lethal outcome to the treated animal [71,72]. Surviving mice developed EBOV-specific antibodies, suggesting that *in vivo* suppression of virus replication by favipiravir allows the host to assemble a virus-specific adaptive immune response to tackle the infection [71].

However, the favipiravir therapeutic effect is hardly beneficial at the terminal stage of the disease. The small animal data generates cautious optimism about translating the findings into nonhuman primate trials. Prophylactic and complement therapeutic treatment with favipiravir against EBOV on nonhuman primates successfully inhibited viral production and extended survival time in a concentration-dependent manner [83]. Another pharmacokinetic study of favipiravir in nonhuman primates against two filoviruses (EBOV and Marburg) also supported the antiviral effect of favipiravir compared with untreated animals. Reduced levels of viral RNA and extended survival time was observed in EBOV-infected nonhuman primates, whereas 83% of the MARV-infected nonhuman primates survived upon treatment with favipiravir [84]. In 2014, a proof-of-concept trial of favipiravir in patients with EVD in Guinea was initiated, in which patients received favipiravir along with standardized care [85]. Data on the safety and effectiveness of favipiravir in reducing mortality and viral load were collected. The study reported a lower fatality rate among patients with low to moderate viral load compared with the control group. The findings were supported by FPV trials in Sierra Leone and different parts of Guinea [15,86].

To study the effect of favipiravir on COVID-19, favipiravir was approved for the treatment of COVID-19 disease on February 15, 2020 in China. A pilot study of a nonrandomized control trial at The Third People’s Hospital of Shenzhen reported significantly better treatment effects in terms of disease progression and viral clearance compared with lopinavir/ritonavir treatment [87]. A different randomized clinical trial was conducted at three hospitals in China (Zhongnam Hospital of Wuhan University, Leishenshan Hospital, The Third People’s Hospital of Hubei Province) to compare the efficacy and safety of favipiravir and arbidol to treat COVID-19 patients [88]. The trial recruited a total of 240 patients and followed up from Feb 20, 2020 to Mar 12, 2020. Patients in the experimental group received various doses of 1600-2400 mg of favipiravir and compared with patients treated with other multiple antiviral drugs: kaletra, oseltamivir, and hydroxychloroquine. Favipiravir-treated patients were found to have a higher clinical recovery rate and more effectively reduced incidence of fever and cough which manifested as mild and moderate symptoms of COVID-19 where during this initial stage, SARS-CoV-2 multiplies and binds to angiotensin-converting enzyme 2 (ACE2) receptor on human cells [7,8]. Other clinical trials of favipiravir monotherapy or combination drug therapy are currently ongoing in China and Thailand to further evaluate the efficacy and safety of favipiravir for the treatment of COVID-19 disease. There is still insufficient data to determine the mechanism of action of favipiravir for the treatment of patients diagnosed with COVID-19.

#### 3.1.4. Chloroquine antimalarial drug with a new effect on COVID-19

In contrast to remdesivir and favipiravir, chloroquine (CQ) is an antimalarial drug considered as one of the drug candidates that exhibit good inhibitory effects on SARS-CoV-2 at the cellular level [62]. Chloroquine is a 9-aminoquinoline that was synthesized in 1934 as an effective substitute for natural quinine used against malaria (Figure 7) [89–91]. Chloroquine has been widely used to treat other human diseases such as amoebiosis, autoimmune diseases, and a wide range of bacterial, fungal, and viral infections [92–96]. Studies have also reported its versatile antiviral activity against RNA viruses as diverse as the rabies virus, poliovirus, HIV, hepatitis viruses, influenza viruses and Ebola virus [97–102].

**Figure 7.**
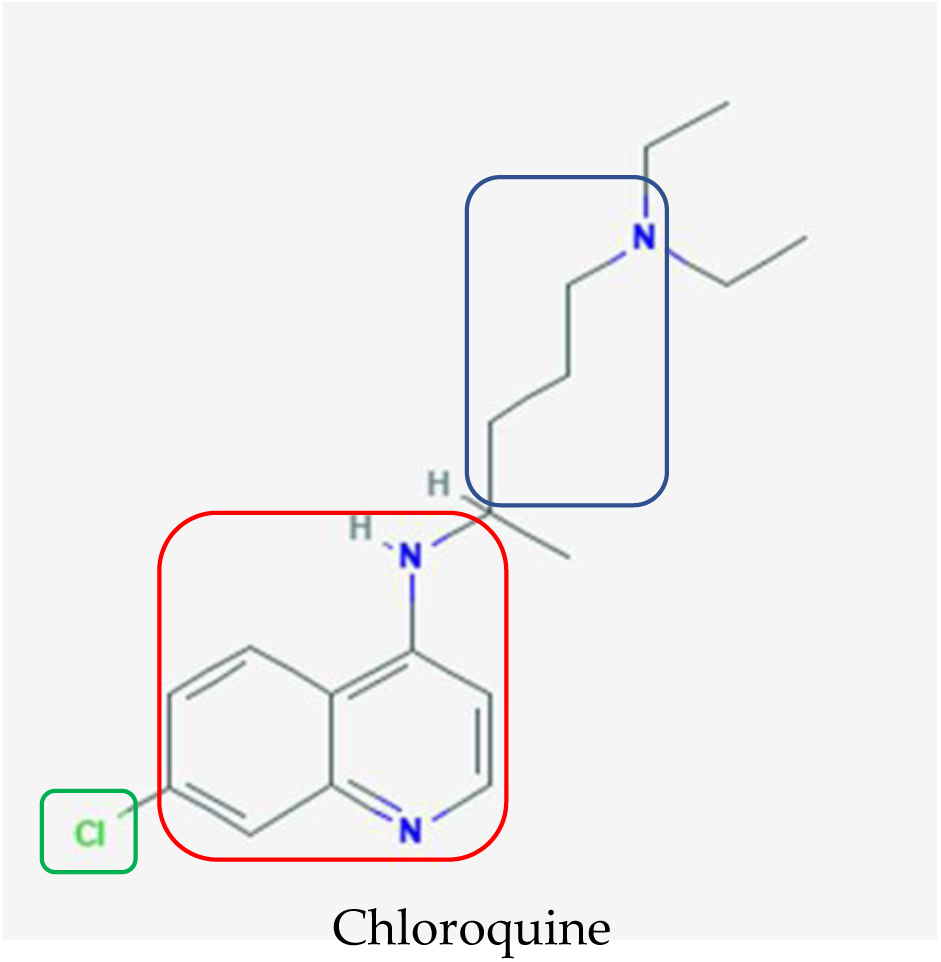
Chemical structure of chloroquine, which was initially used as an anti-malarial agent [103]. The terminal nitrogens and spacer (blue box) were designed to impart parasite resistance whereas the 4-amino quinolone nucleus (red box) and small electron withdrawing group (green box) are pivotal for binding to hematin and halting the formation of hemozoin, respectively, when chloroquine is administered to treat malaria.

Chloroquine is a weak base that accumulates in low-pH organelles, such as endosomes, Golgi vesicles, and lysosomes and interfere with their acidification. This hampers the low-pH-dependent steps of protein degradation and viral replication, including fusion and uncoating [96]. Additionally, it alters the glycosylation of the cellular receptors of viruses, which may affect viral binding [104,105].

The potential activity of chloroquine against coronaviruses has been demonstrated in different *in vitro* studies. Chloroquine successfully inhibited viral replication of HCoV-229E, SARS-CoV, MERS-CoV, and EBOV in various cell lines [104,106–109]. Animal studies, on the other hand, have revealed mixed results. Treatment with chloroquine showed no significant protection against SARS-CoV and EBOV with reports of high toxicity in mouse and hamster models [110,111]. However, other studies have reported positive results against HCoV-OC43 and EBOV when treated with chloroquine [109,112]. The contradict results in animal studies could be owing to the range of doses tested, whereby higher doses could be necessary to produce consistently positive results. However, this may lead to a poor outcome owing to an increase in drug related toxicity. Furthermore, chloroquine may be more effective as a prophylactic treatment owing to its activity during the early stages of a viral cycle, during which it establishes residence in the host through replication of SARS-CoV-2 during the incubation period in patients in the initial stage of the disease, which is a mild condition [7].

Recently, Wang *et al*. [62] reported that the antiviral drugs remdesivir and chloroquine were effective in preventing replication of a clinical isolate of SARS-CoV-2. A clinical trial from more than 100 patients also demonstrated that chloroquine phosphate was superior to the control treatment for inhibiting the exacerbation of pneumonia, improving lung imaging findings, promoting a virus negative conversion and shortening the disease course, which are symptoms during the severe stage of illness in COVID-19 [9]. However, the data should be carefully considered before drawing definitive conclusions, because no other results have been published to support this trial. There are a number of clinical trials for the treatment of COVID-19 using CQ registered in the Chinese Clinical Trial Registry (ChiCTR2000029939, ChiCTR2000029935, ChiCTR2000029899, ChiCTR2000029898, ChiCTR2000029868, ChiCTR2000029837, ChiCTR2000029826, ChiCTR2000029803, ChiCTR2000029762, ChiCTR2000029761, ChiCTR2000029760, ChiCTR2000029741, ChiCTR2000029740, ChiCTR2000029609, ChiCTR2000029559, ChiCTR2000029542) [113,114]. The requests to conduct these clinical trial have been approved and the findings from chloroquine might explore and investigate the mechanism of action of chloroquine to inhibit SARS-CoV-2.

### 3.2. Alternative and complementary medicine used for the treatment of COVID-19

#### 3.2.1. Traditional Chinese medicine can suppress SARS-CoV-2 in vitro

Complementary medicine has also been used to fight this pandemic disease as an alternative medication. One of the complementary or alternative medicines that have been used is *Shuang Huang Lian Kou Fu Ye*. It was officially used on 23 January 2020 by Beijing Administration of Traditional Chinese Medicine [115–117]. It is a Traditional Chinese Medicine (TCM) comprised of three medicinal plants; 375g of *Lonicera japonica*, 375 g of *Scutellaria baicalensis*, and 750 g of *Forsythia suspense* according to Pharmacopoeia of the People’s Republic of China [118]. It is often widely prescribed by Chinese practitioners for the treatment of high fever, coma, and respiratory diseases [119]. According to Professor Zhao Zhi Li from Shanghai Traditional Chinese Medicine University, *Scutellaria baicalensis* is acknowledged as a middle class medicine and *Forsythia suspense* as a low class medicine recorded in Shennong’s Classic of Materia Medica from Han dynasty according to its ranking for medicinal value with high class or A level as the most valued in terms of pharmacology, whereas *Lonicera japonica* has been used as a medicinal plant for the treatment of diseases since the Southern and Northern Dynasty in “Mingyi Bielu” [120,121]. There is a total of 12 companies that manufacture this antidote and one of the manufacturers, Hayao Company’s *Shuang Huang Lian Kou Fu Ye* was categorized as an “A level” list drug, which is the best quality of drugs in TCM according to the categorization in 2019 National health insurance project in China [115].

This complementary medicine was sold out after the announcement made by Wuhan Institute of Virology through articles from Shanghai Institutes for Biological Sciences, CAS, who claimed that this medicine can suppress the SARS-CoV-2 in a cell culture according to a study in collaboration with Shanghai Institute of Materia Medica [122]. However, this TCM was not included in the guideline launched by National Health Commission of the People’s Republic of China for prevention of COVID-19 which have been updated to version 7 and known as Guidelines of Diagnosis and Treatment for COVID-19 version 4 report in China for prevention of COVID-19, as reported by China News [115,123]. This is because the findings were limited to an initial laboratory phase and insufficient data was available to confirm that this TCM can suppress SARS-CoV-2. A clinical trial is still essential to verify its efficacy. As a result, Shanghai Public Health Clinical Center and Wuhan Tongji Hospital have initiated a clinical trial on *Shuang Huang Lian Kou Fu Ye* [124]. Although there is still insufficient scientific evidence that this TCM can be used to control COVID-19, it still widely used and the Beijing Administration of Traditional Chinese Medicine claims that it is one of the TCM that can be used to prevent COVID-19. Because both SARS-CoV and SARS-CoV-2 are from the coronaviruses family, the strategies used for the treatment of SARS could be relevant for COVID-19 [125]. Furthermore, the medicinal plants that in this TCM are known for their antiviral function and have been utilized to alleviate influenza and restrain SARS coronavirus.

All the medicinal plants in this TCM have similar common characteristics, which include heat-clearing, drying dampness and detoxification [126] and antiviral activity against influenza and SARS [127]. For example, *Lonicera japonica* was widely used to control the SARS coronavirus in 2003 according to State Administration of TCM of China [123]. In 2009, *Lonicera japonica* was the second most used medicinal plant for treating influenza A virus subtype H1N1 according to a statistics report from Beijing Youan Hospital of Capital Medical University [128]. The antiviral activity might be owing to the presence of unique compounds such as chlorogenic acid, baicalin, and forsythoside A in *Lonicera japonica, Scutellaria baicalensis*, and *Forsythia suspense* respectively as shown in Figure 8.

**Figure 8.**
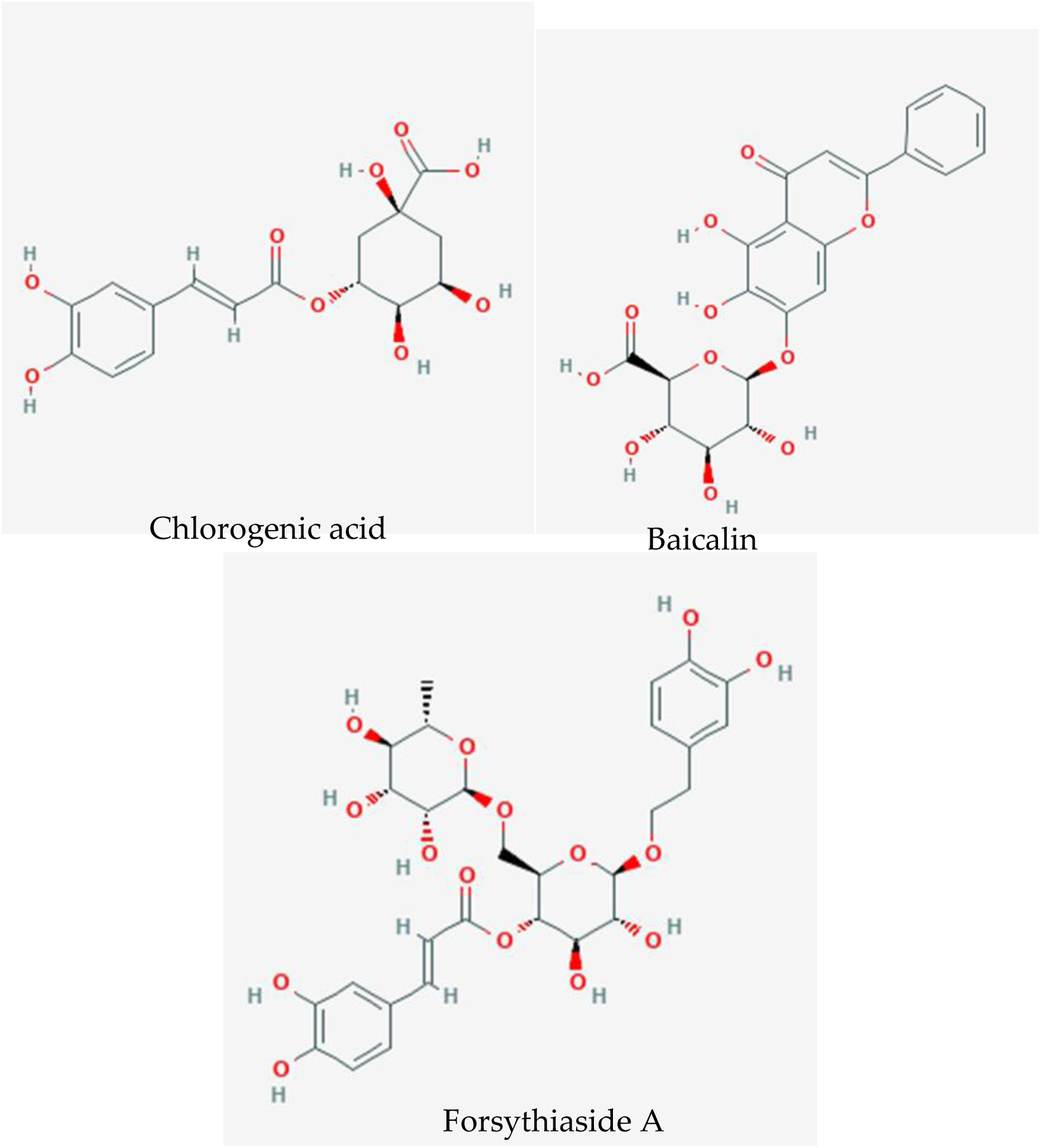
Chemical constituents of *Shuang Huang Lian Kou Fu Ye*, which responsible for the antiviral mechanism of action: Chlorogenic acid [135], baicalin [136]), and forsythiaside A [137].

Chlorogenic acid was abundant in *Lonicera japonica* and has been reported to attenuate the death rate of influenza-virus-infected mice during the late stage of the infectious cycle through down-regulation of nucleoprotein protein expression [129]. Furthermore, chlorogenic acid play a role as neuraminidase blocker to inhibit H1N1 and H3N2 from releasing newly formed virus particles from infected cells. Baicalin, a flavone glycoside found in *Scutellaria baicalensis*, also act as a neuraminidase blocker by activating the JAK/STAT-1 signaling pathway by inducing IFN-γ production in human CD4+ and CD8+ T cells and NK cells and by attenuating miR-146a, a pivotal key in the replication of H1N1 and H3N2, by targeting the TNF-receptor associated factor 6 (TRAF6) [130–133]. Furthermore, *Forsythia suspense* also displayed antiviral activity, which was attributed to the presence of the unique compound forsythoside A [134]. The treatment of influenza with this chemical constituent resulted in a reduction of the viral titers of different influenza virus subtypes in cell cultures owing to a decrease in the Matrix protein 1 protein of influenza and blocked the budding process of the virion formation, which limited the spreading of the virus. These chemical constituents, chlorogenic acid, baicalin, and forsythiaside A, share a common characteristic of being antiviral against influenza; however, their antiviral activity against coronaviruses requires further investigation for further clarification.

### 3.3. Traditional Chinese Medicine recommended by Chinese CDC according to severity of clinical symptoms

Based on the Guidelines of Diagnosis and Treatment for COVID-19 Version 5 by the National Health Commission (NHC) of the People’s Republic of China on 8 February 2020, three TCMs have been used depending on the severity of the condition of COVID-19 and symptom differentiation [123,138].

#### 3.3.1. Treatment of mild clinical symptoms

For patients that showed mild clinical symptoms such as a dry cough, fatigue, chest tightness, nausea, mild cold fever, or no fever, the TCM consisted of 15g of *Atractylodes lancea*, 6g of *Pericarpium Citri Reticulatae*, 10g of *Magnolia officinalis*, 9g of *Agastache rugosa*, 6g of *Amomum tsaoko Crevost et Lemarie*, 9g of *Chinese ephedra*, 10g of *Notopterygium incisum*, 9g of *Zingiber officinale* and 10g of *Areca catechu* were comply [139]. This decoction is a combination of the prescribed TCM *Bu Huan Jin Zheng Qi San* and *Da Yuan Yin* adapted from a medical encyclopedia “Gu Jin Yi Tong Da Quan” chapter 76 and Wu Youke in a book titled On Epidemic Diseases (Wen Yi Lun) respectively from Dynasty Ming [140,141]. According to Shin [142], all the ingredients were categorized based on a method known as a system of chief, deputy and assistant in TCM philosophy or theory. *Atractylodes lancea* is the “chief” ingredient whereas *Agastache rugosa, Pericarpium Citri Reticulatae* and *Magnolia officinalis* act as “deputy” ingredients and *Zingiber officinale* and *Areca catechu* are responsible as “assistant” ingredients to provide better synergistic effect among the herbs for treating patients in best method [143]. Atractylon (Figure 9) is a chemical compound present in the chief ingredient, *Atractylodes lancea*, which attenuated the influenza A virus within 5 days through the TLR7 signaling pathway by upregulating the Toll-like receptor 7 (TLR7), MyD88, tumor necrosis factor receptor-associated factor 6, and IFN-β mRNA expression in the lung tissue of mice infected with influenza A virus [144].

**Figure 9.**
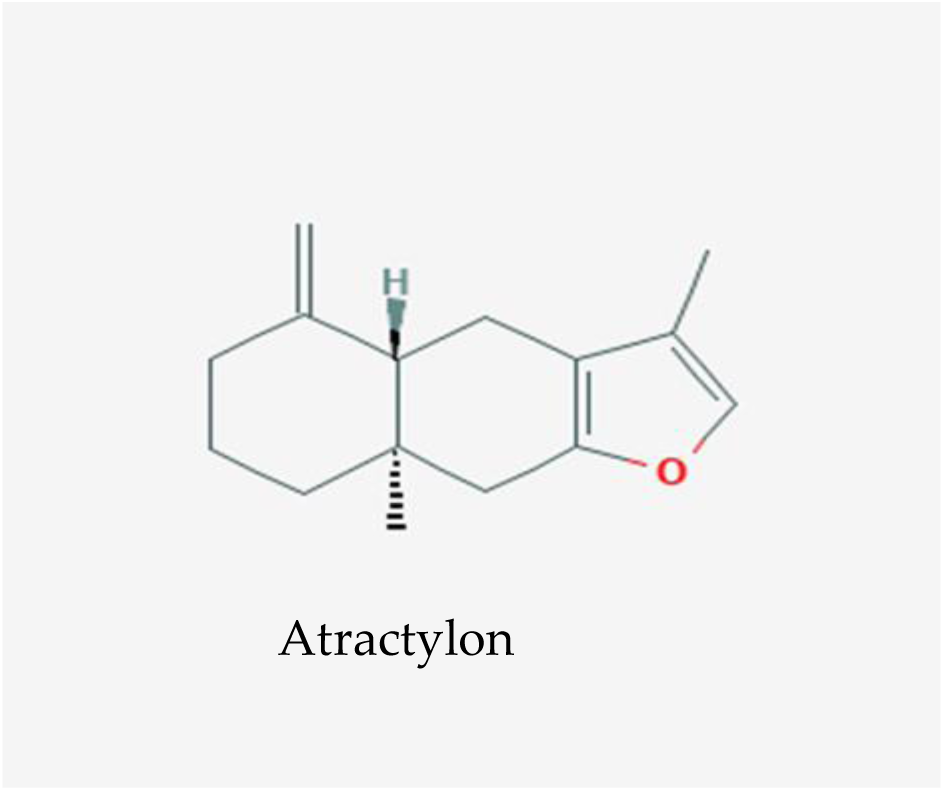
One of the main compounds, atractylon found in the TCM decoction used to treat patients with mild symptoms of COVID-19 [145].

The inhibition of coronaviruses by the synergistic effect of this TCM could be similar in action for inhibition of influenza through triggering of the TLR7 signaling pathway. However, further studies are needed to prove the mechanism of action of this decoction on COVID-19.

#### 3.3.2. Treatment of mild to severe clinical symptoms

Recently, *Qing Fei Pai Du Tang* has been applied for the treatment of patients with any clinical symptoms of COVID-19 ranging from mild to severe cases (incubation period of SARS-CoV-2, viral multiplication and extra-pulmonary systemic hyperinflammation syndrome) [7–9] and even used as a preventative medicine for this disease. 1102 of 1261 confirmed cases in 10 Chinese provinces were reported to be cured and discharged after the treatment with this TCM [146]. Similarly, the Chinese government has reported that from 108 patients diagnosed with mild COVID-19 cases, the number of cases that evolved from mild to severe was approximately 10% when administered Western medicine alone compared with approximately 4.1% when integrated Chinese and Western medicine treatment was used [123]. *Qing Fei Pai Du Tang* is comprised of 21 medicinal plants; 9g of *Chinese ephedra*, 6g of *Glycyrrhiza uralensis*, 9g of *Prunus armeniaca*, 15 to 30g of Gypsum, 9g of *Ramulus cinnamomi*, 9g of *Alisma plantago-aquatica*, 9g of *Polyporus umbellatus*, 9g of *Atractylodes macrocephala*, 15g of *Wolfiporia extensa*, 16g of *Bupleurum chinense*, 6g of *Scutellaria baicalensis*, 9g of *Pinelliae tuber*, 9g of *Zingiber officinale*, 9g of *Aster tataricus*, 9g of *Tussilago farfara*, 9g of *Iris domestica*, 6g of *Asarum*, 12g of *Dioscorea oppositifolia*, 6g of *Trifoliate orange*, 6g of *Pericarpium Citri Reticulatae* and 9g of *Agastache rugosa*. This TCM is a combination of four combinations of well-known prescribed classic TCM from Treatise on Cold Damage Diseases, which are *Ma Xing Shi Gan Tang, She Gan Ma Huang tang, Xiao Cai Hu Tang* and *Wu Ling San* [147].

*Ma Xing Shi Gan Tang* (MXSGT) is known for antipyretic effects and is commonly used to treat pneumonia, influenza, and other respiratory diseases [148]. A systematic review found that the combination of MXSGT with western medicine significantly increased the effective rate of treatment to treat pneumonia (P<0.00001) [149]. The efficacy of this TCM showed significant improvement (P<0.05) on day 7 of consumption of the decoction and was effective and safe for the treatment of community-acquired pneumonia [148]. This effectiveness might be mediated by β2-adrenoceptors in pigs on bronchial smooth muscle to inhibit neutrophil from entering the respiratory airway and can block acetyl-cholinergic and histaminergic receptor-induced bronchial contraction in rats and finally reduce neutrophilic inflammation [150,151]. Furthermore, it plays roles in decreasing IL-4, IL-8, and TNF-α, yet increase IFN-γ in a COPD rat model [152]. In addition to its effectiveness against acute airway system ailments, this decoction also regulate pathogenesis of influenza virus A in infected RAW264.7 cells through attenuation of LC3, the autophagy marker protein [153].

A classic decoction used in China and Japan, *She Gan Ma Huang tang* (SGMHT) inhibits mast cells from releasing substances during inflammation, regulates the viscera’s function, and promotes the apoptosis of eosinophils [154]. It has been reported that mRNA expression levels of Th2 cytokines were decreased and associated with Th1 cytokines upregulation and direct attenuation of the pulmonary edema and suppression of the NF-kB pathway through two herbs (*Aster tataricus* and *Belamcanda chinensis*) from a modified version of SGMHT [151]. In contrast to the other TCMs mentioned above, the other two TCM formulas (*Xiao Cai Hu Tang* and *Wu Ling San*) function differently and did not exhibit function against acute airway obstruction. *Xiao Cai Hu Tang* (XCHT) is known as a TCM for liver treatment, particularly chronic hepatitis B. This TCM modulates STAT3 expression and indirectly suppresses the hepatitis B virus according to western blot analyses and real time PCR results [155]. The synergistic effect of this TCM for the treatment of pneumonia is unknown and requires full elucidation in the future. *Wu Ling San* WLS) has been used for the treatment of impairments of the regulation of body fluid homeostasis in China, Japan, and Korea [156]. WLS might affect the signal transduction pathway such as NF-kB, MAPKs and HO-1 to demonstrate anti-inflammatory effects like MXSGT to treat pneumonia or respiratory diseases in lipopolysaccharide stimulated macrophages [157].

The derived formulation of *Qing Fei Pai Du Tang* from the four combination of these classic TCM showed its ability to reduce the symptoms of COVID-19 patients by restoring the normal body temperature in 94.6 % of 112 patients and stopping coughing in 80.6 % of 214 patients [158]. As a result, this TCM have been listed as one of the treatment option by the National Health Commission (NHC) of the People’s Republic of China in Guidelines of Diagnosis and Treatment for COVID-19 Version 7 [123,138]. The effectiveness of *Qing Fei Pai Du Tang* can reach 97.78% with 1102 patients been cured from 1261 patients from 10 districts in China until 13 March 2020 [123,159]. Until now, none of the cases became severe from mild condition after the consumption of *Qing Fei Pai Du Tang*.

#### 3.3.3. Treatment of severe clinical symptoms

*Xue Bi Jing Injection*, an established TCM in China, has been recommended for severe symptoms of COVID-19 patients [123,138]. Unlike other drugs that undergo a conventional clinical trial phase, it proceeded from bedside to bench and finally back to bedside before approval was obtained from China Food and Drug Administration in 2004 (Chinese Society of Critical Care Medicine, 2015)[160]. Consequently, the annual consumption of this TCM in China for the treatment of pneumonia and other diseases has reached 800000 [160]. *Xue Bi Jing Injection* consist of numerous compounds, as illustrated in Figure 10, which are extracted from five medicinal plants; *Radix Salviae Miltiorrhiae, Rhizoma Chuanxiong, Flos Carthami, Angelica Sinensis* and *Radix Paeoniae Rubra* [161].

**Figure 10.**
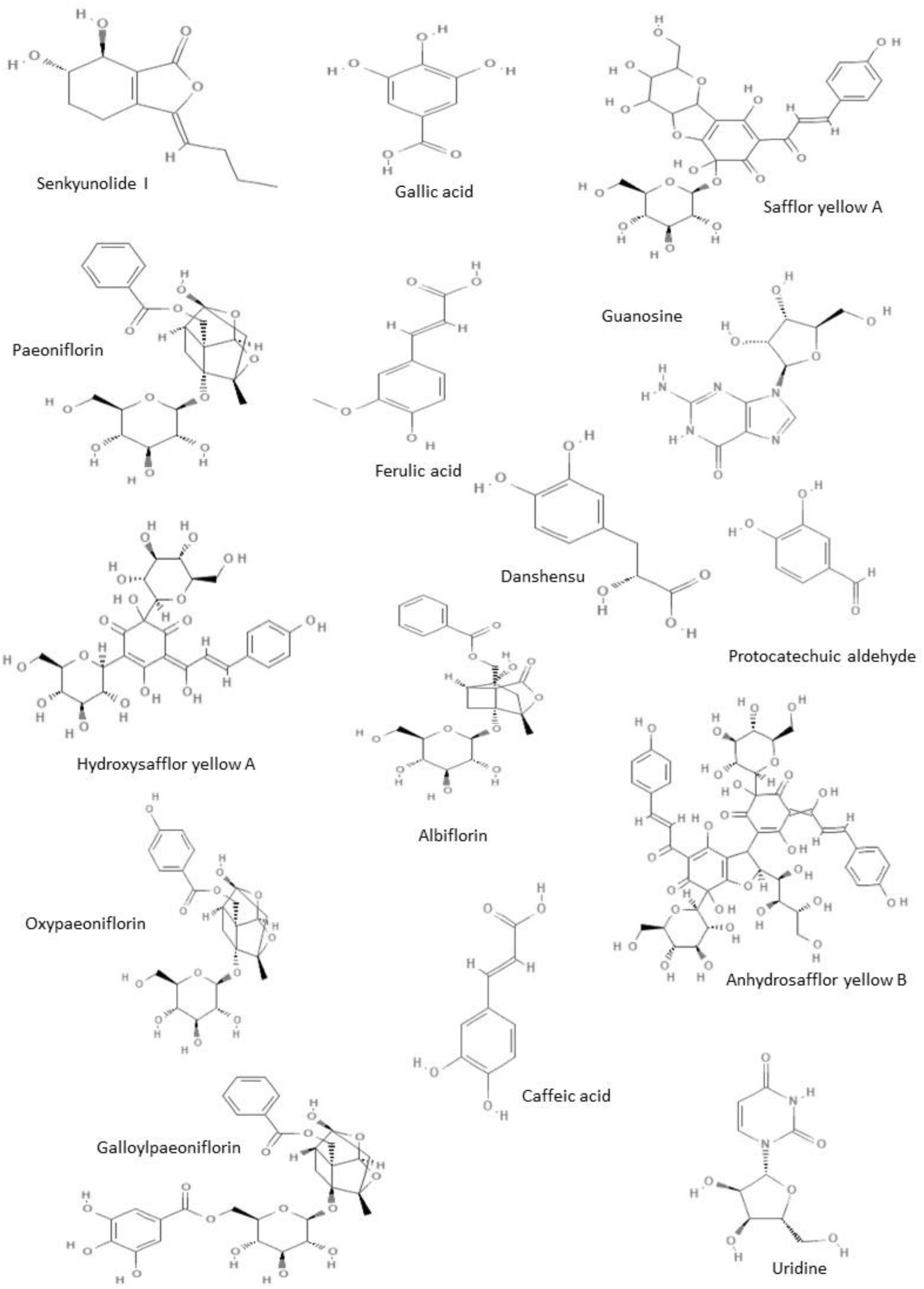
15 chemical constituents in a *Xue Bi Jing Injection*. Senkyunolide I [162], safflor yellow A [163], paeoniflorin [164], ferulic acid [165], galloylpaeoniflorin [166], anhydrosafflor yellow B [167], oxypaeoniflorin [168], caffeic acid [169], albiflorin [170], uridine [171], gallic acid [172], guanosine [173], danshensu [174], protocatechuic aldehyde [175], and hydroxysafflor yellow A [176].

Initially, it was developed for activating blood circulation to remove blood stasis, cooling the blood, and clearing the toxic heat [160]. However, this TCM has been used to fight SARS-CoV-2 because of its effectiveness to treat severe pneumonia, the severe stage of COVID-19 [9], by significantly reducing mortality by approximately 15.9% and elevating the improvement of the pneumonia severity index by approximately 60.8% [160]. The *Xue Bi Jing Injection* relieved or reduced severe pneumonia by triggering the inflammation pathway through downregulation of TNF-α, IL-6 and IL-8 on the 3rd, 7th and 14th day after treatment, although it did not significantly influence the release of leptin [177]. This suggest that an anti-endotoxin effect was deployed by halting the release of TNF- α, IL-6 and IL-8, endogenic inflammatory mediators. As a result, blocking the development of a systemic inflammatory response syndrome occurred through the disruption of the inflammation vicious cycle [177]. This mechanism of action of the *Xue Bi Jing Injection* reduces the severity of pneumonia in COVID-19 patients and lowers the side effects on the organ functions.

These results indicate that TCMs can be used as complementary medicine for the treatment of patients during this pandemic disease; however, the mechanism of action of these TCMs still require further investigation.

## 4. Materials and Methods

A systematic scoping review was conducted from 1 January 2020 until 18 March 2020 by including papers that were published and not published before 18 March 2020 and reported drug repurposing and traditional Chinese medicine (TCM) as treatment options for COVID-19 used in patients from different countries to reduce publication bias, increase the comprehensiveness and timeliness of the review and foster a balanced picture of available evidence [178]. The review was performed according to criteria using Preferred Reporting Items for Systematic Reviews (PRISMA) statement [179]. The list of publications was obtained from listed databases and search engines: PubMed (https://www.ncbi.nlm.nih.gov/pubmed/), Science Direct (https://www.sciencedirect.com/), Google Scholar(https://scholar.google.com/) and Semantic Scholar (https://www.semanticscholar.org/), whereas grey literature search such as guidelines, theses and dissertations, research and committee reports, government reports, conference papers, and abstracts were obtained from WHO, U.S National Library of Medicine, Chinese Clinical Trial Registry (http://www.chictr.org.cn/abouten.aspx), China Centre for disease control and prevention, China Important Conference Papers Database, China Dissertation Database, and online official news websites. Most of the grey literatures were from China because the virus originated from China and China provides more data to tackle this disease. Different combinations of the following keywords: NCov-2019, COVID-19, 2019-nCoV, SARS-CoV-2, 2019 novel coronavirus, Wuhan virus, drug repurposing, medication, treatment, traditional Chinese medicine, alternative and complementary medicine were used in the literature search through a three-level search strategy based on standardized descriptors defined by the Medical Subject Headings algorithm. A secondary search was based on screening of the reference list of all the relevant studies identified in the direct search. The entire potentially relevant studies were evaluated after title screening to exclude irrelevant information. Only studies that reported the repurposing of drugs and TCMs for COVID-19 and contained information about the structure of the chemical constituents, *in vivo* or *in vitro* studies, case reports, treatment of patients diagnosed with COVID-19, and molecular mechanisms were extracted and assessed.

## 5. Conclusions

The treatment of COVID-19 with repurposed drugs is dependent on the ability of the drug to inhibit the proliferation by binding to the enzyme active sites, viral chain termination, and triggering of molecular pathways. For instance, ritonavir and lopinavir are believed to bind at the enzyme active site and are used together to inhibit the replication of SARS-CoV-2 because lopinavir alone is susceptible to oxidation metabolism by the cytochrome P450 enzyme, whereas the addition of ritonavir can inhibit P450 3A4 enzyme activity. Therefore, the combination of ritonavir and lopinavir are essential to maintain the drug efficacy for the treatment of COVID-19. However, the mechanism of oseltamivir combined with kaletra remains unclear and needs further experimental validation. Therefore, the combination of ritonavir and lopinavir are essential to maintain the drug efficacy for the treatment of COVID-19 during their function in preventing the binding to the SARS-CoV 3CL^pro^ enzyme site, which is responsible for the replication of SARS-CoV-2 glycoprotein. Furthermore, remdesivir and favipiravir attenuated replication of SARS-CoV-2 through premature termination of RNA transcription. Chloroquine shows a potential antiviral effect *in vitro*, however, there is a lack of *in vivo* data to support therapeutic treatment for SARS-CoV-2 infection. In contrast to the 6 drugs mentioned above, the 4 TCMs discussed in this review were initially used to treat influenza and SARS by acting as neuraminidase blockers and triggering the inflammation pathway such as blocking the release of endogenic inflammatory mediators, TNF- α, IL-6, and IL-8 to halt the inflammation mechanism of action instead of RNA chain termination. New drugs that are effective for the treatment COVID-19 are still in development or in clinical trials. In this review we provide a framework for better understanding of the mechanism of action of repurposed drugs and TCMs and their involvement in the molecular pathway for inhibiting viral replication such as SARS or MERS. However, there are limitation to our findings because there is still a lack of concrete evidence for the mechanisms of action of the drugs and their curative effect on COVID-19. Further experimental validation is needed to provide more concrete evidence. We hope this will provide insights into the molecular pathway and lead to the development of effective drugs for the treatment of COVID-19 in the future. However, further experimental validation is needed to provide more concrete evidence.

## Data Availability

All data is provided in the manuscript.

## Acknowledgments

We would like to thank the Director General of Health Malaysia for his permission to publish this article

## Ethical Declaration

Ethical clearance is exempted, as approved by the Medical Research and Ethics Committee (MREC), Ministry of Health Malaysia (NMRR-20-347-53302) because no personal information or samples from human subjects are used in this systematic scoping review.

## References

1. Gorbalenya, A.E.; Baker, S.C.; Baric, R.S.; Gulyaeva, A.A.; Lauber, C.; I.A., S.; A.M., L.; Penzar, D.; Samborskiy, D.; Groot, R.J.; et al. The species Severe acute respiratory syndrome-related coronavirus: classifying 2019-nCoV and naming it SARS-CoV-2. Nat. Microbiol. 2020, 5, 536–544, doi:10.1038/s41564-020-0695-z.

2. Naming the coronavirus disease (COVID-19) and the virus that causes it Available online: https://www.who.int/emergencies/diseases/novel-coronavirus-2019/technical-guidance/naming-the-coronavirus-disease-(covid-2019)-and-the-virus-that-causes-it (accessed on Apr 21, 2020).

3. Coronavirus disease 2019 (COVID-19): Situation Report - 66 Available online: https://www.who.int/docs/default-source/coronaviruse/situation-reports/20200326-sitrep-66-covid-19.pdf?sfvrsn=81b94e61_2.

4. Zhou, P.; Yang, X.-L.; Wang, X.-G.; Hu, B.; Zhang, L.; Zhang, W.; Si, H.-R.; Zhu, Y.; Li, B.; Huang, C.-L.; et al. A pneumonia outbreak associated with a new coronavirus of probable bat origin. Nature 2020, 579, 270–273, doi:10.1038/s41586-020-2012-7.

5. Huang, C.; Wang, Y.; Li, X.; Ren, L.; Zhao, J.; Hu, Y.; Zhang, L.; Fan, G.; Xu, J.; Gu, X.; et al. Clinical features of patients infected with 2019 novel coronavirus in Wuhan, China. The Lancet 2020, 395, 497–506, doi:10.1016/S0140-6736(20)30183-5.

6. Wang, D.; Hu, B.; Hu, C.; Zhu, F.; Liu, X.; Zhang, J.; Wang, B.; Xiang, H.; Cheng, Z.; Xiong, Y.; et al. Clinical Characteristics of 138 Hospitalized Patients With 2019 Novel Coronavirus–Infected Pneumonia in Wuhan, China. JAMA 2020, 323, 1061–1069, doi:10.1001/jama.2020.1585.

7. Wan, Y.; Shang, J.; Graham, R.; Baric, R.S.; Li, F. Receptor Recognition by the Novel Coronavirus from Wuhan: an Analysis Based on Decade-Long Structural Studies of SARS Coronavirus. J. Virol. 2020, 94, doi:10.1128/JVI.00127-20.

8. Siddiqi, H.K.; Mehra, M.R. COVID-19 Illness in Native and Immunosuppressed States: A Clinical-Therapeutic Staging Proposal. J. Heart Lung Transplant. 2020, S105324982031473X, doi:10.1016/j.healun.2020.03.012.

9. Qin, C.; Zhou, L.; Hu, Z.; Zhang, S.; Yang, S.; Tao, Y.; Xie, C.; Ma, K.; Shang, K.; Wang, W.; et al. Dysregulation of immune response in patients with COVID-19 in Wuhan, China. Clin. Infect. Dis. 2020, ciaa248, doi:10.1093/cid/ciaa248.

10. van Doremalen, N.; Bushmaker, T.; Morris, D.H.; Holbrook, M.G.; Gamble, A.; Williamson, B.N.; Tamin, A.; Harcourt, J.L.; Thornburg, N.J.; Gerber, S.I.; et al. Aerosol and Surface Stability of SARS-CoV-2 as Compared with SARS-CoV-1. N. Engl. J. Med. 2020, 382, 1564–1567, doi:10.1056/NEJMc2004973.

11. Shereen, M.A.; Khan, S.; Kazmi, A.; Bashir, N.; Siddique, R. COVID-19 infection: Origin, transmission, and characteristics of human coronaviruses. J. Adv. Res. 2020, 24, 91–98, doi:10.1016/j.jare.2020.03.005.

12. Cascella, M.; Rajnik, M.; Cuomo, A.; Dulebohn, S.C.; Di Napoli, R. Features, Evaluation and Treatment Coronavirus (COVID-19). In StatPearls; StatPearls Publishing: Treasure Island (FL), 2020.

13. Rothe, C.; Schunk, M.; Sothmann, P.; Bretzel, G.; Froeschl, G.; Wallrauch, C.; Zimmer, T.; Thiel, V.; Janke, C.; Guggemos, W.; et al. Transmission of 2019-nCoV Infection from an Asymptomatic Contact in Germany. N. Engl. J. Med. 2020, 382, 970–971, doi:10.1056/NEJMc2001468.

14. Yu, P.; Zhu, J.; Zhang, Z.; Han, Y. A Familial Cluster of Infection Associated With the 2019 Novel Coronavirus Indicating Possible Person-to-Person Transmission During the Incubation Period. J. Infect. Dis. 2020, jiaa077, doi:10.1093/infdis/jiaa077.

15. Bai, C.Q.; Mu, J.S.; Kargbo, D.; Song, Y.B.; Niu, W.K.; Nie, W.M.; Jiang, J.F. Clinical and Virological characteristics of Ebola virus disease patients treated with Favipiravir (T-705)-Sierra Leone, 2014. Clin. Infect. Dis. 2016, 63, 1288–1294.

16. Callaway, E. Coronavirus vaccines: five key questions as trials begin. Nature 2020, 579, 481.

17. Weingartl, H.; Czub, M.; Czub, S.; Neufeld, J.; Marszal, P.; Gren, J.; Smith, G.; Jones, S.; Proulx, R.; Deschambault, Y.; et al. Immunization with modified vaccinia virus Ankara-based recombinant vaccine against severe acute respiratory syndrome is associated with enhanced hepatitis in ferrets. J. Virol. 2004, 78, 12672–12676.

18. Xu, J.; Zhao, S.; Teng, T.; Abdalla, A.E.; Zhu, W.; Xie, L.; Wang, Y.; Guo, X. Systematic Comparison of Two Animal-to-Human Transmitted Human Coronaviruses: SARS-CoV-2 and SARS-CoV. Viruses 2020, 12, doi:10.3390/v12020244.

19. Lu, H. Drug treatment options for the 2019-new coronavirus (2019-nCoV). Biosci. Trends 2020, 14, 69–71, doi:10.5582/bst.2020.01020.

20. WHO to launch multinational trial to jumpstart search for coronavirus drugs. STAT 2020.

21. Analogue-based drug discovery; Fischer, J., Ganellin, C.R., Eds.; Wiley-VCH: Weinheim, 2006; ISBN 978–3–527–31257–3.

22. Yao, T.-T.; Qian, J.-D.; Zhu, W.-Y.; Wang, Y.; Wang, G.-Q. A Systematic Review of Lopinavir Therapy for SARS Coronavirus and MERS Coronavirus-A Possible Reference for Coronavirus Disease-19 Treatment Option. J. Med. Virol. 2020, jmv.25729, doi:10.1002/jmv.25729.

23. NORVIR® Label; Abbott Laboratories, 2011;

24. Wishart, D.S.; Feunang, Y.D.; Guo, A.C.; Lo, E.J.; Marcu, A.; Grant, J.R.; Sajed, T.; Johnson, D.; Li, C.; Sayeeda, Z.; et al. DrugBank 5.0: a major update to the DrugBank database for 2018. Nucleic Acids Res. 2018, 46, D1074–D1082, doi:10.1093/nar/gkx1037.

25. PubChem Ritonavir Available online: https://pubchem.ncbi.nlm.nih.gov/compound/392622 (accessed on Apr 21, 2020).

26. PubChem Lopinavir Available online: https://pubchem.ncbi.nlm.nih.gov/compound/92727 (accessed on Apr 21, 2020).

27. Kempf, D.J. 8.15 - Ritonavir and Lopinavir/Ritonavir. In Comprehensive Medicinal Chemistry II; Taylor, J.B., Triggle, D.J., Eds.; Elsevier: Oxford, 2007; pp. 187–197 ISBN 978–0–08–045044–5.

28. Cuevas, J.M.; Geller, R.; Garijo, R.; López-Aldeguer, J.; Sanjuán, R. Extremely High Mutation Rate of HIV-1 In Vivo. PLOS Biol. 2015, 13, e1002251, doi:10.1371/journal.pbio.1002251.

29. Rock, B.M.; Hengel, S.M.; Rock, D.A.; Wienkers, L.C.; Kunze, K.L. Characterization of ritonavir-mediated inactivation of cytochrome P450 3A4. Mol. Pharmacol. 2014, 86, 665–674, doi:10.1124/mol.114.094862.

30. Lv, Z.; Chu, Y.; Wang, Y. HIV protease inhibitors: A review of molecular selectivity and toxicity. HIVAIDS - Res. Palliat. Care 2015, 7, 95–104, doi:10.2147/HIV.S79956.

31. Van Waterschoot, R.A.B.; Ter Heine, R.; Wagenaar, E.; Van Der Kruijssen, C.M.M.; Rooswinkel, R.W.; Huitema, A.D.R.; Beijnen, J.H.; Schinkel, A.H. Effects of cytochrome P450 3A (CYP3A) and the drug transporters P-glycoprotein (MDR1/ABCB1) and MRP2 (ABCC2) on the pharmacokinetics of lopinavir. Br. J. Pharmacol. 2010, 160, 1224–1233, doi:10.1111/j.1476-5381.2010.00759.x.

32. Woman, 74, recovers from virus after “Thai cocktail” Available online: https://www.bangkokpost.com/thailand/general/1860329/woman-74-recovers-from-virus-after-thai-cocktail (accessed on Apr 21, 2020).

33. Harrison, C. Coronavirus puts drug repurposing on the fast track. Nat. Biotechnol. 2020, doi:10.1038/d41587-020-00003-1.

34. Coronavirus patient “recovers after being treated with a HIV drug” | Daily Mail Online Available online: https://www.dailymail.co.uk/health/article-8077889/Coronavirus-patient-recovers-treated-HIV-drug.html (accessed on Apr 21, 2020).

35. Nukoolkarn, V.; Lee, V.S.; Malaisree, M.; Aruksakulwong, O.; Hannongbua, S. Molecular dynamic simulations analysis of ritronavir and lopinavir as SARS-CoV 3CLpro inhibitors. J. Theor. Biol. 2008, 254, 861–867, doi:10.1016/j.jtbi.2008.07.030.

36. Zhang, X.W.; Yap, Y.L. Old drugs as lead compounds for a new disease? Binding analysis of SARS coronavirus main proteinase with HIV, psychotic and parasite drugs. Bioorg. Med. Chem. 2004, 12, 2517–2521, doi:10.1016/j.bmc.2004.03.035.

37. Yamamoto, N.; Yang, R.; Yoshinaka, Y.; Amari, S.; Nakano, T.; Cinatl, J.; Rabenau, H.; Doerr, H.W.; Hunsmann, G.; Otaka, A.; et al. HIV protease inhibitor nelfinavir inhibits replication of SARS-associated coronavirus. Biochem. Biophys. Res. Commun. 2004, 318, 719–725, doi:10.1016/j.bbrc.2004.04.083.

38. Chu, C.M.; Cheng, V.C.C.; Hung, I.F.N.; Wong, M.M.L.; Chan, K.H.; Chan, K.S.; Kao, R.Y.T.; Poon, L.L.M.; Wong, C.L.P.; Guan, Y.; et al. Role of lopinavir/ritonavir in the treatment of SARS: Initial virological and clinical findings. Thorax 2004, 59, 252–256, doi:10.1136/thorax.2003.012658.

39. Sheahan, T.P.; Sims, A.C.; Leist, S.R.; Schäfer, A.; Won, J.; Brown, A.J.; Montgomery, S.A.; Hogg, A.; Babusis, D.; Clarke, M.O.; et al. Comparative therapeutic efficacy of remdesivir and combination lopinavir, ritonavir, and interferon beta against MERS-CoV. Nat. Commun. 2020, 11, doi:10.1038/s41467-019-13940-6.

40. Hoffmann, M.; Kleine-Weber, H.; Schroeder, S.; Mü, M.A.; Drosten, C.; Pö, S.; Krü, N.; Herrler, T.; Erichsen, S.; Schiergens, T.S.; et al. SARS-CoV-2 Cell Entry Depends on ACE2 and TMPRSS2 and Is Blocked by a Clinically Proven Protease Inhibitor Article SARS-CoV-2 Cell Entry Depends on ACE2 and TMPRSS2 and Is Blocked by a Clinically Proven Protease Inhibitor. Cell 2020, 181, 1–10, doi:10.1016/j.cell.2020.02.052.

41. Casadevall, A.; Pirofski, L.-A. The convalescent sera option for containing COVID-19. J. Clin. Invest. 2020, 130, doi:10.1172/JCI138003.

42. Bloomberg China Seeks Plasma From Recovered Patients as Virus Treatment - Bloomberg Available online: https://www.bloomberg.com/news/articles/2020-02-14/china-seeks-plasma-from-recovered-patients-for-coronavirus-cure.

43. Jones, M.; Hama, R.; DelMar, C. Oseltamivir for influenza. The Lancet 2014, 386, 1133–1134.

44. McNicholl, I.R.; McNicholl, J.J. Neuraminidase inhibitors: zanamivir and oseltamivir. Ann. Pharmacother. 2001, 35, 57–70, doi:10.1345/aph.10118.

45. Hoffmann, M.; Kleine-Weber, H.; Schroeder, S.; Krüger, N.; Herrler, T.; Erichsen, S.; Schiergens, T.S.; Herrler, G.; Wu, N.-H.; Nitsche, A.; et al. SARS-CoV-2 Cell Entry Depends on ACE2 and TMPRSS2 and Is Blocked by a Clinically Proven Protease Inhibitor. Cell 2020, 181, 271–280.e8, doi:10.1016/j.cell.2020.02.052.

46. Genentech, I. Tamiflu (oseltamivir phosphate) capsules label; 2010;

47. PubChem Oseltamivir Available online: https://pubchem.ncbi.nlm.nih.gov/compound/65028 (accessed on Apr 21, 2020).

48. Tan, E.L.C.; Ooi, E.E.; Lin, C.Y.; Tan, H.C.; Ling, A.E.; Lim, B.; Stanton, L.W. Inhibition of SARS Coronavirus Infection in Vitro with Clinically Approved Antiviral Drugs. Emerg. Infect. Dis. 2004, 10, 581–586, doi:10.3201/eid1004.030458.

49. Al-Tawfiq, J.A.; Momattin, H.; Dib, J.; Memish, Z.A. Ribavirin and interferon therapy in patients infected with the Middle East respiratory syndrome coronavirus: An observational study. Int. J. Infect. Dis. 2014, 20, 42–46, doi:10.1016/j.ijid.2013.12.003.

50. Tchesnokov, E.P.; Feng, J.Y.; Porter, D.P.; Gotte, M. Mechanism of inhibition of Ebola virus RNA-dependent RNA polymerase by remdesivir. Viruses 2019, 11, 326.

51. Lo, M.K. GS-5734 and its parent nucleoside analog inhibit Filo-, Pneumo-, and Paramyxoviruses. Sci Rep 2017, 7, 43395.

52. Sheahan, T.P.; Sims, A.C.; Graham, R.L.; Menachery, V.D.; Gralinski, L.E.; Case, J.B.; Leist, S.R.; Pyrc, K.; Feng, J.Y.; Trantcheva, I.; et al. Broad-spectrum antiviral GS-5734 inhibits both epidemic and zoonotic coronaviruses. Sci. Transl. Med. 2017, 9, eaal3653, doi:10.1126/scitranslmed.aal3653.

53. Warren, T.K. Therapeutic efficacy of the small molecule GS-5734 against Ebola virus in rhesus monkeys. Nature 2016, 531, 381–385.

54. Dörnemann, J. First newborn baby to receive experimental therapies survives Ebola virus disease. J Infect Dis 2017, 215, 171–174.

55. Mulangu, S.; Dodd, L.E.; Davey, R.T.; Tshiani Mbaya, O.; Proschan, M.; Mukadi, D.; Lusakibanza Manzo, M.; Nzolo, D.; Tshomba Oloma, A.; Ibanda, A.; et al. A Randomized, Controlled Trial of Ebola Virus Disease Therapeutics. N. Engl. J. Med. 2019, 381, 2293–2303, doi:10.1056/NEJMoa1910993.

56. PubChem Remdesivir Available online: https://pubchem.ncbi.nlm.nih.gov/compound/121304016 (accessed on Apr 21, 2020).

57. Gordon, C.J.; Tchesnokov, E.P.; Feng, J.Y.; Porter, D.P.; Götte, M. The antiviral compound remdesivir potently inhibits RNA-dependent RNA polymerase from Middle East respiratory syndrome coronavirus. J. Biol. Chem. 2020, 295, 4773–4779, doi:10.1074/jbc.AC120.013056.

58. Brown, A.J.; Won, J.J.; Graham, R.L.; Dinnon, K.H.; Sims, A.C.; Feng, J.Y.; Cihlar, T.; Denison, M.R.; Baric, R.S.; Sheahan, T.P. Broad spectrum antiviral remdesivir inhibits human endemic and zoonotic deltacoronaviruses with a highly divergent RNA dependent RNA polymerase. Antiviral Res. 2019, 169, 104541, doi:10.1016/j.antiviral.2019.104541.

59. Agostini, M.L.; Andres, E.L.; Sims, A.C.; Graham, R.L.; Sheahan, T.P.; Lu, X. Coronavirus susceptibility to the antiviral remdesivir (GS-5734) is mediated by the viral polymerase and the proofreading exoribonuclease. mBio 2018, 2018;9, 1128 00221–18.

60. de Wit, E.; Feldmann, F.; Cronin, J.; Jordan, R.; Okumura, A.; Thomas, T.; Scott, D.; Cihlar, T.; Feldmann, H. Prophylactic and therapeutic remdesivir (GS-5734) treatment in the rhesus macaque model of MERS-CoV infection. Proc. Natl. Acad. Sci. 2020, 117, 6771–6776, doi:10.1073/pnas.1922083117.

61. Sheahan, T.P.; Sims, A.C.; Leist, S.R.; Schäfer, A.; Won, J.; Brown, A.J.; Montgomery, S.A.; Hogg, A.; Babusis, D.; Clarke, M.O.; et al. Comparative therapeutic efficacy of remdesivir and combination lopinavir, ritonavir, and interferon beta against MERS-CoV. Nat. Commun. 2020, 11, 222, doi:10.1038/s41467-019-13940-6.

62. Wang, M.; Cao, R.; Zhang, L.; Yang, X.; Liu, J.; Xu, M. Remdesivir and chloroquine effectively inhibit the recently emerged novel coronavirus (2019-nCoV) in vitro. Cell Res. 2020, 30, 269–271.

63. Holshue, M.L.; DeBolt, C.; Lindquist, S.; Lofy, K.H.; Wiesman, J.; Bruce, H. Washington State 2019-nCoV Case Investigation Team. N. Engl. J. Med. 2020, 382, 929–936.

64. A Trial of Remdesivir in Adults With Severe COVID-19 Available online: https://clinicaltrials.gov/ct2/show/NCT04257656.

65. Furuta, Y.; Takahashi, K.; Fukuda, Y.; Kuno, M.; Kamiyama, T.; Kozaki, K.; Nomura, N.; Egawa, H.; Minami, S.; Watanabe, Y.; et al. In vitro and in vivo activities of anti-influenza virus compound T-705. Antimicrob Agents Chemother 2002, 46, 977–981.

66. Furuta, Y.; Gowen, B.B.; Takahashi, K.; Shiraki, K.; Smee, D.F.; Barnard, D.L. Favipiravir (T-705), a novel viral RNA polymerase inhibitor. Antivir. Res 2013, 100, 446–454, doi:10.1016/j.antiviral.2013.09.015.

67. Rocha-Pereira, J.; Jochmans, D.; Dallmeier, K.; Leyssen, P.; Nascimento, M.S.J.; Neyts, J. Favipiravir (T-705) inhibits in vitro norovirus replication. Biochem Biophys Res Commun 2012, 424, 777–780, doi:10.1016/j.

68. Zmurko, J.; Marques, R.E.; Schols, D.; Verbeken, E.; Kaptein, S.J.F.; Neyts, J. The Viral Polymerase Inhibitor 7-Deaza-2’-C-Methyladenosine Is a Potent Inhibitor of In Vitro Zika Virus Replication and Delays Disease Progression in a Robust Mouse Infection Model. PLoS Negl. Trop. Dis. 2016, 10, e0004695, doi:10.1371/journal.pntd.0004695.

69. de los Santos, T.; de Avila Botton, S.; Weiblen, R.; Grubman, M.J. The Leader Proteinase of Foot-and-Mouth Disease Virus Inhibits the Induction of Beta Interferon mRNA and Blocks the Host Innate Immune Response. J. Virol. 2006, 80, 1906–1914, doi:10.1128/JVI.80.4.1906-1914.2006.

70. Yamada, K.; Noguchi, K.; Komeno, T.; Furuta, Y.; Nishizono, A. Efficacy of Favipiravir (T-705) in Rabies Postexposure Prophylaxis. J. Infect. Dis. 2016, 213, 1253–1261, doi:10.1093/infdis/jiv586.

71. Oestereich, L.; Lüdtke, A.; Wurr, S.; Rieger, T.; Muñoz-Fontela, C.; Günther, S. Successful treatment of advanced Ebola virus infection with T-705 (favipiravir) in a small animal model. Antiviral Res. 2014, 105, 17–21.

72. Smither, S.J.; Eastaugh, L.S.; Steward, J.A.; Nelson, M.; Lenk, R.P.; Lever, M.S. Post-exposure efficacy of Oral T-705 (Favipiravir) against inhalational Ebola virus infection in a mouse model. Antiviral Res. 2014, 104, 153–155, doi:10.1016/j.antiviral.2014.01.012.

73. Favipiravir | C5H4FN3O2 - PubChem Available online: https://pubchem.ncbi.nlm.nih.gov/compound/492405#section=2D-Structure (accessed on Apr 21, 2020).

74. Sangawa, H.; Komeno, T.; Nishikawa, H.; Yoshida, A.; Takahashi, K.; Nomura, N.; Furuta, Y. Mechanism of action of T-705 ribosyl triphosphate against influenza virus RNA polymerase. Antimicrob Agents Chemother 2013, 57, 5202–5208.

75. Abdelnabi, R.; Morais, A.T.S. de; Leyssen, P.; Imbert, I.; Beaucourt, S.; Blanc, H.; Froeyen, M.; Vignuzzi, M.; Canard, B.; Neyts, J.; et al. Understanding the Mechanism of the Broad-Spectrum Antiviral Activity of Favipiravir (T-705): Key Role of the F1 Motif of the Viral Polymerase. J. Virol. 2017, 91, doi:10.1128/JVI.00487-17.

76. Smee, D.F.; Hurst, B.L.; Egawa, H.; Takahashi, K.; Kadota, T.; Furuta, Y. Intracellular metabolism of favipiravir (T-705) in uninfected and influenza A (H5N1) virus-infected cells. J Antimicrob Chemother 2009, 64, 741–746.

77. Arias, A.; Thorne, L.; Goodfellow, I. Favipiravir elicits antiviral mutagenesis during virus replication in vivo. eLife 2014, 3, e03679, doi:10.7554/eLife.03679.

78. Furuta, Y.; Takahashi, K.; Kuno-Maekawa, M.; Sangawa, H.; Uehara, S.; Kozaki, K.; Nomura, N.; Egawa, H.; Shiraki, K. Mechanism of Action of T-705 against Influenza Virus. Antimicrob. Agents Chemother. 2005, 49, 981–986, doi:10.1128/AAC.49.3.981-986.2005.

79. Jin, Z.; Smith, L.K.; Rajwanshi, V.K.; Kim, B.; Deval, J. The Ambiguous Base-Pairing and High Substrate Efficiency of T-705 (Favipiravir) Ribofuranosyl 5′-Triphosphate towards Influenza A Virus Polymerase. PLoS ONE 2013, 8, e68347, doi:10.1371/journal.pone.0068347.

80. Naesens, L.; Guddat, L.W.; Keough, D.T.; van Kuilenburg, A.B.P.; Meijer, J.; Vande Voorde, J.; Balzarini, J. Role of Human Hypoxanthine Guanine Phosphoribosyltransferase in Activation of the Antiviral Agent T-705 (Favipiravir). Mol. Pharmacol. 2013, 84, 615–629, doi:10.1124/mol.113.087247.

81. Baranovich, T.; Wong, S.-S.; Armstrong, J.; Marjuki, H.; Webby, R.J.; Webster, R.G.; Govorkova, E.A. T-705 (favipiravir) induces lethal mutagenesis in influenza A H1N1 viruses in vitro. J Virol 2013, 87, 3741–3751, doi:10.1128/JVI.02346-12.

82. de Ávila, A.I.; Gallego, I.; Soria, M.E.; Gregori, J.; Quer, J.; Esteban, J.I.; Rice, C.M.; Domingo, E.; Perales, C. Lethal Mutagenesis of Hepatitis C Virus Induced by Favipiravir. PLOS ONE 2016, 11, e0164691, doi:10.1371/journal.pone.0164691.

83. Guedj, J.; Piorkowski, G.; Jacquot, F.; Madelain, V.; Nguyen, T.H.T.; Rodallec, A. Antiviral efficacy of favipiravir against Ebola virus: A translational study in cynomolgus macaques. PLoS Med 2018, 15, 1002535.

84. Bixler, S.L.; Bocan, T.M.; Wells, J.; Wetzel, K.S.; Van Tongeren, S.A.; Dong, L.; Garza, N.L.; Donnelly, G.; Cazares, L.H.; Nuss, J.; et al. Efficacy of favipiravir (T-705) in nonhuman primates infected with Ebola virus or Marburg virus. Antiviral Res. 2018, 151, 97–104, doi:10.1016/j.antiviral.2017.12.021.

85. Sissoko, D.; Laouenan, C.; Folkesson, E.; M’Lebing, A.-B.; Beavogui, A.-H.; Baize, S.; Camara, A.-M.; Maes, P.; Shepherd, S.; Danel, C.; et al. Experimental Treatment with Favipiravir for Ebola Virus Disease (the JIKI Trial): A Historically Controlled, Single-Arm Proof-of-Concept Trial in Guinea. PLoS Med. 2016, 13, e1001967, doi:10.1371/journal.pmed.1001967.

86. Kerber Laboratory Findings, Compassionate Use of Favipiravir, and Outcome in Patients With Ebola Virus Disease, Guinea, 2015-A. Retrosp. Obs. Study J. Infect. Dis. 220, 195–202.

87. Cai, Q.; Yang, M.; Liu, D.; Chen, J.; Shu, D.; Xia, J.; Liao, X.; Gu, Y.; Cai, Q.; Yang, Y.; et al. Experimental Treatment with Favipiravir for COVID-19: An Open-Label Control Study. Engineering 2020, S2095809920300631, doi:10.1016/j.eng.2020.03.007.

88. Chen, C.; Zhang, Y.; Huang, J.; Yin, P.; Cheng, Z.; Wu, J.; Chen, S.; Zhang, Y.; Chen, B.; Lu, M.; et al. Favipiravir versus Arbidol for COVID-19: A Randomized Clinical Trial; Infectious Diseases (except HIV/AIDS), 2020;

89. Bruce-Chwatt, L.J.; Black, R.H.; Canfield, C.J.; Clyde, D.F.; Peters, W.; Wernsdorfer, W.H.; Organization, W.H. Chemotherapy of malaria; World Health Organization, 1986; ISBN 978–92–4–140127–2.

90. Winzeler, E.A. Malaria research in the post-genomic era. Nature 2008, 455, 751–6.

91. Parhizgar, A.R.; Tahghighi, A. Introducing new antimalarial analogues of chloroquine and amodiaquine: a narrative review. Iran J Med Sci 2017, 42, 115–28.

92. Lee, S.J.; Silverman, E.; Bargman, J.M. The role of antimalarial agents in the treatment of SLE and lupus nephritis. Nat Rev Nephrol 2011, 7, 718–29, doi:10.1038/nrneph.2011.150.

93. Raoult, D.; Drancourt, M.; Vestris, G. Bactericidal effect of doxycycline associated with lysosomotropic agents on Coxiella burnetii in P388D1 cells. Antimicrob Agents Chemother 1990, 34, 1512–4, doi:10.1128/aac.34.8.1512.

94. Raoult, D.; Houpikian, P.; Tissot, D.H.; Riss, J.M.; Arditi-Djiane, J.; Brouqui, P. Treatment of Q fever endocarditis: comparison of 2 regimens containing doxycycline and ofloxacin or hydroxychloroquine. Arch Intern Med 1999, 159, 167–73, doi:10.1001/archinte.159.2.167.

95. Boulos, A.; Rolain, J.M.; Raoult, D. Antibiotic susceptibility of Tropheryma whipplei in MRC5 cells. Antimicrob Agents Chemother 2004, 48, 747–52.

96. Rolain, J.M.; Colson, P.; Raoult, D. Recycling of chloroquine and its hydroxyl analogue to face bacterial, fungal and viral infection in the 21st century. Int J Antimicrob Agents 2007, 30, 297–308.

97. Boelaert, P., JR; J, S.; K. The potential place of chloroquine in the treatment of HIV-1-infected patients. J Clin Virol 2001, 20, 137–40.

98. Dowall, S.D.; Bosworth, A.; Watson, R.; Bewley, K.; Taylor, I.; Rayner, E. Chloroquine inhibited Ebola virus replication in vitro but failed to protect against infection and disease in the in vivo guinea pig model. J Gen Virol 2015, 96, 3484–92.

99. Kronenberger, P.; Vrijsen, R.; Boeyé, A. Chloroquine induces empty capsid formation during poliovirus eclipse. J Virol 1991, 65, 7008–11.

100. Vigerust, D.J.; McCullers, J.A. Chloroquine is effective against influenza A virus in vitro but not in vivo. Influenza Other Respir. Viruses 2007, 1, 189–192, doi:10.1111/j.1750-2659.2007.00027.x.

101. Devaux, C.A.; Rolain, J.-M.; Colson, P.; Raoult, D. New insights on the antiviral effects of chloroquine against coronavirus: what to expect for COVID-19? Int. J. Antimicrob. Agents 2020, 105938, doi:10.1016/j.ijantimicag.2020.105938.

102. Mizui, T.; Yamashina, S.; Tanida, I.; Takei, Y.; Ueno, T.; Sakamoto, N. Inhibition of hepatitis C virus replication by chloroquine targeting virus-associated autophagy. J Gastroenterol 2010, 45, 195–203.

103. PubChem Chloroquine Available online: https://pubchem.ncbi.nlm.nih.gov/compound/2719 (accessed on Apr 21, 2020).

104. Savarino, A.; Boelaert, C., JR; A, M.; G, C.; R. Effects of chloroquine on viral infections: an old drug against today’s diseases? Lancet Infect Dis 2003, 3, 722–727.

105. Vincent, M.J.; Bergeron, E.; Benjannet, S.; Erickson, B.R.; Rollin, P.E.; Ksiazek, T.G.; Seidah, N.G.; Nichol, S.T. Chloroquine is a potent inhibitor of SARS coronavirus infection and spread. Virol. J. 2005, 2, 69, doi:10.1186/1743-422X-2-69.

106. Blau, D.M.; Holmes, K.V. Human Coronavirus HCoV-229E enters susceptible cells via the endocytic pathway. Adv. Exp. Med. Biol. 2001, 494, 193–198.

107. Colson, P.; Rolain, J.-M.; Raoult, D. Chloroquine for the 2019 novel coronavirus SARS-CoV-2. Int. J. Antimicrob. Agents 2020, 55, 105923, doi:10.1016/j.ijantimicag.2020.105923.

108. Johansen, L.M.; Brannan, J.M.; Delos, S.E.; Shoemaker, C.J.; Stossel, A.; Lear, C.; Hoffstrom, B.G.; Dewald, L.E.; Schornberg, K.L.; Scully, C.; et al. FDA-approved selective estrogen receptor modulators inhibit Ebola virus infection. Sci Transl Med 2013, 5, 190 179.

109. Madrid, P.B.; Chopra, S.; Manger, I.D.; Gilfillan, L.; Keepers, T.R.; Shurtleff, A.C.; Green, C.E.; Iyer, L.V.; Dilks, H.H.; Davey, R.A.; et al. A Systematic Screen of FDA-Approved Drugs for Inhibitors of Biological Threat Agents. PLoS ONE 2013, 8, e60579, doi:10.1371/journal.pone.0060579.

110. Barnard, D.L.; Day, C.W.; Bailey, K.; Heiner, M.; Montgomery, R.; Lauridsen, L.; Chan, P.K.; Sidwell, R.W. Evaluation of immunomodulators, interferons and known in vitro SARS-coV inhibitors for inhibition of SARS-coV replication in BALB/c mice. Antivir Chem Chemother 2006, 17, 275–284.

111. Falzarano, D.; Safronetz, D.; Prescott, J.; Marzi, A.; Feldmann, F.; Feldmann, H. Lack of protection against ebola virus from chloroquine in mice and hamsters. Emerg Infect Dis 2015, 21 1065–1067.

112. Keyaerts, E.; Li, S.; Vijgen, L.; Rysman, E.; Verbeeck, J.; Van Ranst, M.; Maes, P. Antiviral activity of chloroquine against human coronavirus OC43 infection in newborn mice. Antimicrob. Agents Chemother. 2009, 53, 3416–3421, doi:10.1128/AAC.01509-08.

113. Chinese Clinical Trial Register (ChiCTR) - The world health organization international clinical trials registered organization registered platform Available online: http://www.chictr.org.cn/enIndex.aspx (accessed on Apr 21, 2020).

114. Kearney, J. Chloroquine as a Potential Treatment and Prevention Measure for the 2019 Novel Coronavirus: A Review; MEDICINE & PHARMACOLOGY, 2020;

115. Morse, J.S.; Lalonde, T.; Xu, S.; Liu, W.R. Learning from the Past: Possible Urgent Prevention and Treatment Options for Severe Acute Respiratory Infections Caused by 2019–nCoV. ChemBioChem 2020, 21, 730–738, doi:10.1002/cbic.202000047.

116. *Pneumonia prevention plan for new coronavirus infection in Beijing Version 1*; Beijing Administration of Traditional Chinese Medicine, 2020;

117. Liu, X. Shuang Huang Lian that “cure everything” sold out Available online: https://m.chinanews.com/wap/detail/zw/cj/2020/02-01/9074862.shtml.

118. Pharmacopoeia Committee of the People’s Republic of China; China Medical Science and Technology Press: Beijing, 2015;

119. Wu, S.; Xu, L.; Liu, H.; Tong, X. Clinical application and dosage of Coptidis Rhizoma. Chin. Clin. Dr. 2015, 43, 92–94.

120. Wang, J.; Wang, L.; Lou, G.H.; Zeng, H.R.; Hu, J.; Huang, Q.W.; Peng, W.; Yang, X.B. Coptidis Rhizoma: a comprehensive review of its traditional uses, botany, phytochemistry, pharmacology and toxicology. Pharm. Biol. 2019, 57, 193–225.

121. Xu, R.Z. Epidemic prevention new science: What are the use of the three medicines in “Shuang Huang Lian” and is it effective? What do the TCM experts say? (translated) Available online: https://www.jfdaily.com/news/detail?id=205513.

122. Xie, X. Shuang Huang Lian and the manufacturer behind it Available online: https://www.jiemian.com/article/3932685.html.

123. Yang, Y.; Islam, M.S.; Wang, J.; Li, Y.; Chen, X. Traditional Chinese Medicine in the Treatment of Patients Infected with 2019-New Coronavirus (SARS-CoV-2): A Review and Perspective. Int. J. Biol. Sci. 2020, 16, 1708–1717.

124. Yue, H.B. Shanghai Institute of Materia Medica finding: Shuang Huang Lian Kou Fu Ye can suppress new coronavirus Available online: http://scitech.people.com.cn/n1/2020/0131/c1007-31566098.html.

125. Chen, Y.; Liu, Q.; Guo, D. Emerging coronaviruses: Genome structure, replication, and pathogenesis. J. Med. Virol. 2020, 92, 418–423.

126. Muluye, R.A.; Bian, Y.H.; Alemu, P.N. Anti-inflammatory and antimicrobial effects of heat-clearing chinese herbs: A current review. J. Tradit. Complement. Med. 2014, 4, 93–98.

127. Zheng Investigation of prescription to treat influenza A virus subtype H1N1 in Beijing Youan hospital. Shanxi Med. J. 2010, 39, 897–898.

128. Shang, X.F.; Pan, H.; Li, M.X.; Miao, X.L.; Ding, H. Lonicera japonica Thunb.:Ethnopharmacology, phytochemistry and pharmacology of an important traditional chinese medicine. J. Ethnopharmacol. 2011, 138, 1–21.

129. Ding, Y.; Cao, Z.Y.; Cao, L.; Ding, G.; Wang, Z.Z.; Xiao, W. Antiviral activity of chlorogenic acid against influenza A (H1N1/H3N2) virus and its inhibition of neuraminidase. Sci. Rep. 2017, 7, 45723.

130. Ding, Y.; Dou, J.; Teng, Z.; Yu, J.; Wang, T.; Lu, N.; Wang, H.; Zhou, C. Antiviral activity of baicalin against influenza A (H1N1/H3N2) virus in cell culture and in mice and its inhibition of neuraminidase. Arch. Virol. 2014, 159, 3269–3278.

131. Chu, M.; Xu, L.; Zhang, M.-B.; Chu, Z.-Y.; Wang, Y.-D. Role of Baicalin in Anti-Influenza Virus A as a Potent Inducer of IFN-Gamma. BioMed Res. Int. 2015, 2015, 263630, doi:10.1155/2015/263630.

132. Zhao, T.T.; Tang, H.L.; Xie, L.; Zheng, Y.; Ma, Z.B.; Sun, Q.; Li, X.F. Scutellaria baicalensis Georgi. (Lamiaceae): a review of its traditional uses, botany, phytochemistry, pharmacology and toxicology. J. Pharm. Pharmacol. 2019, 71, 1353–1369.

133. Li, R.; Wang, L.X. Baicalin inhibits influenza virus A replication via activation of type I IFN signaling by reducing miR–146a. Mol. Med. Rep. 2019, 20, 5041–5049.

134. Law, H.Y.A.; Yang, L.H.C.; Lau, S.Y.A.; Chan, G.C.F. Antiviral effect of forsythoside A from Forsythia suspensa (Thunb.) Vahl fruit against influenza A virus through reduction of viral M1 protein. J. Ethnopharmacol. 2017, 209, 236–247.

135. PubChem Chlorogenic acid Available online: https://pubchem.ncbi.nlm.nih.gov/compound/1794427 (accessed on Apr 21, 2020).

136. PubChem Baicalin Available online: https://pubchem.ncbi.nlm.nih.gov/compound/64982 (accessed on Apr 21, 2020).

137. PubChem Forsythiaside Available online: https://pubchem.ncbi.nlm.nih.gov/compound/5281773 (accessed on Apr 21, 2020).

138. Chinese Centre for disease control and prevention; 2020;

139. Guo, J.L.; Yan, Z.H.; Zhou, L. buhuanjin zheng qi san" aroma awaken the spleen experiment. China J. Tradit. Chin. Med. Pharm. 1989, 4, 25–28.

140. Wang, J.Y.; Ren, J.; Chen, S.B.; Yao, S.Z.; Ma, L.; Liu, Z.D. Simultaneous determination of six components such as mangiferin in Dayuan yin by HPLC. J. Tianjin Univ. Tradit. Chin. Med. 2018, 37, 72–75.

141. Li, X.Y.; Xiong, Z.F.; Gu, D.L. Clinical observation of “buhuanjin zheng qi san” in treatment of chemotherapy-induced diarrhea. Chin. J. Tradit. Med. Sci. Technol. 2018, 25, 426–428.

142. Shin, S.S. Analysis of Agastache Powder to Rectify the Ki Combination for the Formula Science Common Textbook. Herb. Formula Sci. 2013, 21, 16–35.

143. Mu, C.W. Determination of Hesperidin in Buhuanjin Zhengqi Powder by RP-HPLC. J. Liaoning Univ. Tradit. Chin. Med. 2013, 3, 49–50.

144. Cheng, Y.; Mai, J.Y.; Hou, T.L.; Ping, J.; Chen, J.J. Antiviral activities of atractylon from Atractylodis Rhizoma. Mol. Med. Rep. 2016, 14, 3704–3710.

145. PubChem Atractylon Available online: https://pubchem.ncbi.nlm.nih.gov/compound/3080635 (accessed on Apr 21, 2020).

146. Zhang, H.P. TCM set for global resurgence Available online: https://www.globaltimes.cn/content/1180189.shtml.

147. Zhang, Z.J.; Feng, Y.; Wiseman, N.; Mitchell, C.; Feng, Y. Shang Han Lun On Cold Damage Translation Commentaries; Paradigm Press, 2000;

148. Gong, X.F. Efficacy and Safety Evaluation of Treatment of CAP Patients with Lung Phlegm Heat Syndrome by Augmented Maxingshigan Decoction Iontophoresis. Master Thesis, Beijing University of Chinese Medicine, 2018.

149. Li, L.; Lu, F.G.; He, Q.H. Efficacy of Maxing Shigan Decoction combined with Western medicine for pneumonia in children: A systematic review and meta-analysis. J. Integr. Med. 2009, 7;809–813.

150. Kao, S.T.; Yeh, T.J.; Hsieh, C.C.; Shiau, H.B.; Yeh, F.T.; Lin, J.G. The effects of ma-xing-gan-shi-tang on respiratory resistance and airway leukocyte infiltration in asthmatic guinea pigs. Immunopharmacol. Immunotoxicol. 2001, 23, 445–458.

151. Eng, Y.S.; Lee, C.H.; Lee, W.C.; Huang, C.C.; Chang, J.S. Unraveling the Molecular Mechanism of Traditional Chinese Medicine: Formulas Against Acute Airway Viral Infections as Examples. Molecules 2019, 24, 3505, doi:10.3390/molecules24193505.

152. Zhang, W.; Xinyue, Z.; Shao, Y. Changes in the level of cytokine in rats with chronic obstructive pulmonary disease of phlegm heat cumber lung type after treatment of Maxing Shigan decoction. Chin. J. Tissue Eng. Res. 2006, 10, 167–170.

153. Li, L.; Zhang, B.; Lu, F.G.; Cai, L.; Gao, Q.; Hu, J.; He, G.L.; Wu, T. Effect of type A influenza virus on autophagy of lung macrophages and intervention of serum of Maxing Shigan decoction. Chin. Pharmacol. Bull. 2019, 35, 878–882.

154. Zhu, P.Y. The Literature and Theory’s Research of Shenggan Mahuang Decoction’s Application on Lung Disease, Master Thesis (Jinan University, 2014.

155. Chen, S.F.; Wang, Z.L.; Wan, S.C.; Huang, H.; Liang, H.Q. Effect of modified Xiaochaihu decoction-containing serum on HepG2.2.15 cells via the JAK2/STAT3 signaling pathway. Mol. Med. Rep. 2017, 16, 7416–7422.

156. Ahn, Y.M.; Cho, K.W.; Kang, D.G.; Lee, H.S. Oryeongsan (Wulingsan), a traditional Chinese herbal medicine, induces natriuresis and diuresis along with an inhibition of the renin-angiotensin-aldosterone system in rats. J. Ethnopharmacol. 2012, 141, 780–785.

157. Oh, Y.C.; Jeong, Y.H.; Ha, J.H.; Cho, W.K.; Ma, J.Y. Oryeongsan inhibits LPS-induced production of inflammatory mediators via blockade of the NF-kappaB, MAPK pathways and leads to HO-1 induction in macrophage cells. BMC Complement. Altern. Med. 2014, 14, 242.

158. Gu, L.P. TCM shows good effects in COVID-19 treatment: official Available online: http://www.ecns.cn/news/2020-02-17/detail-ifztrmvi9823509.shtml.

159. Zhang, X.L.; Zhang, Z.H. The effect of Qingfei Detox Decoction is optimized by a combination of multiple classics of exogenous fever Available online: http://www.bjnews.com.cn/health/2020/02/17/690804.html.

160. Zhang, N.T.; Cheng, C.; Olaleye, O.E.; Sun, Y.; Li, L.; Huang, Y.H.; Du, F.F.; Yang, J.L.; Wang, F.Q.; Shi, Y.H.; et al. Pharmacokinetics-Based Identification of Potential Therapeutic Phthalides from XueBiJing, a Chinese Herbal Injection Used in Sepsis Management. Drug Metab. Dispos. Biol. Fate Chem. 2018, 46, 823–834.

161. Gong, P.; Lu, Z.D.; Xing, J.; Wang, N.; Zhang, Y. Traditional Chinese Medicine Xuebijing Treatment Is Associated with Decreased Mortality Risk of Patients with Moderate Paraquat Poisoning. PLOS ONE 2015, 10, 0130508.

162. PubChem Senkyunolide I Available online: https://pubchem.ncbi.nlm.nih.gov/compound/11521428 (accessed on Apr 21, 2020).

163. PubChem 3,4,6,9-Tetrahydroxy-2-(hydroxymethyl)-8-[(E)-3-(4-hydroxyphenyl)prop-2-enoyl]-6-[(2S,3R,4S,5S,6R)-3,4,5-trihydroxy-6-(hydroxymethyl)oxan-2-yl]oxy-3,4,4a,9b-tetrahydro-2H-pyrano[3,2-b][1]benzofuran-7-one Available online: https://pubchem.ncbi.nlm.nih.gov/compound/71463725 (accessed on Apr 21, 2020).

164. Paeoniflorin | C23H28O11 - PubChem Available online: https://pubchem.ncbi.nlm.nih.gov/compound/442534#section=2D-Structure (accessed on Apr 21, 2020).

165. PubChem Ferulic acid Available online: https://pubchem.ncbi.nlm.nih.gov/compound/445858 (accessed on Apr 21, 2020).

166. PubChem Galloylpaeoniflorin Available online: https://pubchem.ncbi.nlm.nih.gov/compound/46882879 (accessed on Apr 21, 2020).

167. PubChem Anhydrosafflor yellow B Available online: https://pubchem.ncbi.nlm.nih.gov/compound/102240413 (accessed on Apr 21, 2020).

168. PubChem Oxypaeoniflorin Available online: https://pubchem.ncbi.nlm.nih.gov/compound/21631105 (accessed on Apr 21, 2020).

169. PubChem Caffeic acid Available online: https://pubchem.ncbi.nlm.nih.gov/compound/689043 (accessed on Apr 21, 2020).

170. PubChem Albiflorin Available online: https://pubchem.ncbi.nlm.nih.gov/compound/24868421 (accessed on Apr 21, 2020).

171. PubChem Uridine Available online: https://pubchem.ncbi.nlm.nih.gov/compound/6029 (accessed on Apr 21, 2020).

172. PubChem Gallic acid Available online: https://pubchem.ncbi.nlm.nih.gov/compound/370 (accessed on Apr 21, 2020).

173. PubChem Guanosine Available online: https://pubchem.ncbi.nlm.nih.gov/compound/135398635 (accessed on Apr 21, 2020).

174. PubChem Danshensu Available online: https://pubchem.ncbi.nlm.nih.gov/compound/11600642 (accessed on Apr 21, 2020).

175. PubChem 3,4-Dihydroxybenzaldehyde Available online: https://pubchem.ncbi.nlm.nih.gov/compound/8768 (accessed on Apr 21, 2020).

176. PubChem Safflomin A Available online: https://pubchem.ncbi.nlm.nih.gov/compound/6443665 (accessed on Apr 21, 2020).

177. Qi, F.; Liang, Z.X.; She, D.Y.; Yan, G.T.; Chen, L.A. A Clinical Study on the Effects and Mechanism of Xuebijing Injection (血必净注射液) in Severe Pneumonia Patients. J. Tradit. Chin. Med. 2011, 31, 46–49.

178. Paez, A. Grey literature: An important resource in systematic reviews. J. Evid.-Based Med. 2017, 10, 233–240, doi:10.1111/jebm.12265.

179. Moher, D.; Liberati, A.; Tetzlaff, A.; D.G.; Group, P.R.I.S.M.A. Preferred reporting items for systematic reviews and meta-analyses: the PRISMA statement. PLoS Med. 2009, 6, 1000097.

